# Cerebrovascular Reactivity to Hypoxic and Hypercapnic Gas Challenges Using 7T BOLD fMRI

**DOI:** 10.64898/2026.09.11.26362747

**Authors:** S. LoParco, M. Miedema, G.Y. Choi, M. Sayeh, J. Wang, M.H. Boudrias, G.D. Mitsis, R. Hoge

## Abstract

**Background:** Cerebrovascular reactivity (CVR) is a sensitive marker of vascular health, most commonly measured with BOLD fMRI during hypercapnic gas challenges. Hypoxia is a complementary but under-characterized vasoactive stimulus. We characterize a combined hypercapnic-hypoxic gas-challenge paradigm at 7 Tesla BOLD fMRI and its cross-session reproducibility.

**Methods:** Eleven healthy adults were scanned twice at 7 Tesla during an 18-minute multi-echo, multiband BOLD acquisition, receiving fixed-inspired hypercapnic (5% CO_2_) and hypoxic (10% O_2_) gas boluses separated by medical-air recovery periods, with end-tidal gas and peripheral saturation monitoring. Hypercapnic and hypoxic CVR were estimated using three trace-based general linear models — end-tidal gas, peripheral saturation, and Severinghaus-derived arterial saturation — and a finite impulse response model. Cross-session reproducibility was assessed by region-of-interest intraclass correlation.

**Results:** The paradigm produced robust, condition-specific gray-matter BOLD responses that reproduced across sessions. The Severinghaus-derived saturation regressor generalized significantly better than end-tidal and peripheral regressors under cross-session validation and outperformed the finite impulse response model in 58% of gray-matter voxels. Reproducibility across cortical, subcortical, and brainstem atlases was moderate-to-high (median cortical intraclass correlation 0.71).

**Conclusion:** A combined hypercapnic-hypoxic paradigm yields reproducible voxelwise cerebrovascular reactivity maps at 7 Tesla; Severinghaus-derived arterial saturation is the most generalizable regressor for hypoxic reactivity.

## Background

### 1.1 Clinical importance of cerebrovascular reactivity

Cerebrovascular reactivity (CVR) reflects the capacity of the cerebral vasculature to dilate or constrict in response to vasoactive stimuli, and is widely regarded as a sensitive marker of cerebrovascular reserve and vascular health. CVR is altered in healthy aging (Lu et al., 2011), hypertension and vascular aging (Jefferson et al., 2018), stroke and steno-occlusive disease (Mandell et al., 2008), cerebral small-vessel disease (Blair et al., 2016), and a range of neurodegenerative disorders including Alzheimer’s disease and cerebral amyloid angiopathy (Switzer et al., 2020; Yezhuvath et al., 2012). CVR has been proposed both as a diagnostic biomarker (Krishnamurthy et al., 2021; Liu et al., 2019) and as a target for tracking therapeutic intervention, including after surgical revascularization in steno-occlusive disease (McKetton et al., 2019).

### 1.2 Modalities and stimulus paradigms for CVR mapping

Because CVR is defined operationally as the change in a hemodynamic signal per unit change in a vasoactive challenge, both the choice of imaging modality and the choice of stimulus shape what CVR is being measured. Quantitative measurement of cerebral blood flow (CBF) is achievable using arterial spin labeling (ASL), widely regarded as the gold-standard non-invasive MRI sequence for cerebral perfusion. However, ASL suffers from intrinsically low signal-to-noise ratio (SNR), a limitation that is compounded at ultra-high field (7 T) by *B*_0_ and *B*_1_ inhomogeneities that degrade labeling efficiency and readout fidelity (Zuo et al., 2013). The blood-oxygenation-level-dependent (BOLD) signal acquired with fMRI provides an alternative, indirect readout of vasoactive responses, reflecting a composite of changes in CBF, cerebral blood volume, and deoxyhemoglobin content rather than CBF alone; it offers substantially higher SNR per unit time, and has been established as a robust surrogate for mapping cerebrovascular reactivity to controlled vasoactive challenges (Bandettini & Wong, 1997; Kastrup et al., 1999; Liu et al., 2019; Mandell et al., 2008). On the stimulus side, a spectrum of paradigms has been developed: voluntary breath-hold tasks (Bright & Murphy, 2013; Keeling et al., 2025), fixed-inspired gas manipulation in which a known concentration of CO_2_ or O_2_ is delivered via a non-rebreathing or continuous-flow circuit (Bandettini & Wong, 1997; Kastrup et al., 1999), computer-controlled prospective end-tidal targeting using sequential gas delivery (Slessarev et al., 2007) or dynamic end-tidal forcing systems (Wise et al., 2007), pharmacological administration of vasoactive agents such as acetazolamide (Vorstrup et al., 1986), and resting-state methods that exploit spontaneous fluctuations in PCO_2_ as an endogenous probe of vascular responsiveness (Keeling et al., 2025; Prokopiou et al., 2019; Shams et al., 2023; Wise et al., 2004). Transcranial Doppler ultrasound (TCD) offers a complementary, non-imaging modality for CVR characterization based on cerebral blood flow velocity in large basal arteries; while TCD lacks the regional coverage of MRI, it has been used extensively to model dynamic cerebrovascular responses to both spontaneous and manipulated PCO_2_ variations (Mitsis et al., 2004). These paradigms differ in the magnitude, repeatability, and physiological specificity of the resulting PCO_2_ change, and several studies have shown they yield systematically different CVR estimates (Keeling et al., 2025; Liu et al., 2019; Tancredi & Hoge, 2013).

### 1.3 Fixed-inspired versus prospective end-tidal gas administration paradigms

Among these approaches, gas-challenge paradigms in which the inspired gas mixture is precisely controlled — whether by prospective end-tidal targeting or by delivery of a fixed-concentration mixture through a continuous-flow circuit — offer particular advantages over voluntary tasks: they produce large, repeatable changes in arterial blood gas tensions that are largely independent of subject effort and respiration induced motion (Tancredi & Hoge, 2013). The simultaneous capnographic and oximetric monitoring allows the resulting end-tidal traces to be temporally aligned to the BOLD signal for voxelwise CVR mapping (Liu et al., 2019). Within this family, two implementation philosophies sit at opposite ends of a control-versus-accessibility spectrum. Prospective end-tidal targeting — using sequential gas delivery circuits (the RespirAct lineage) (Slessarev et al., 2007) or model-based dynamic end-tidal forcing (Wise et al., 2007) — drives end-tidal PCO_2_ and PO_2_ to a prescribed trajectory on a breath-by-breath basis, decoupling the achieved arterial stimulus from subject-specific ventilatory responses and yielding highly reproducible step-wise blood-gas tensions. Continuous-flow fixed-inspired delivery, in contrast, supplies a known concentration of CO_2_ or O_2_ through an open circuit and allows the participant’s own ventilatory response to set the resulting end-tidal trajectory; it trades the breath-by-breath end-tidal precision of prospective targeting for greater accessibility and substantially simpler apparatus (Tancredi & Hoge, 2013), and is well suited to studies in which compliance with active breathing tasks may be limited. Fixed-inspired continuous-flow delivery has nonetheless served as a workhorse CVR stimulus since the earliest BOLD fMRI experiments (Bandettini & Wong, 1997; Kastrup et al., 1999; Rostrup et al., 1995) and remains widely used, producing vasoactive challenges large and repeatable enough to robustly evoke voxelwise BOLD CVR despite the loss of fine breath-by-breath control over the achieved end-tidal trajectory.

### 1.4 The hypoxic BOLD response and origins of fMRI

Despite the diversity of CVR paradigms, the overwhelming majority deliver a single class of stimulus — hypercapnia — and therefore probe only one dimension of vascular control (Bandettini & Wong, 1997; Bhogal et al., 2015; Blair et al., 2016; Bright & Murphy, 2013; Jefferson et al., 2018; Kastrup et al., 1999; Keeling et al., 2025; Liu et al., 2019; Lu et al., 2011; Mandell et al., 2008; McKetton et al., 2019; Prokopiou et al., 2019; Tancredi & Hoge, 2013; Thomas et al., 2014; Thomas et al., 2013; Yezhuvath et al., 2012). This dominance reflects the fact that the cerebral vasculature is exquisitely sensitive to even small changes in PCO_2_ via a well-characterized CO_2_-mediated vasodilatory pathway, whereas the vascular response to hypoxia is more complex — modulated by ventilatory compensation that can induce concurrent hypocapnic vasoconstriction at mild-to-moderate hypoxic doses, and transitioning to substantial vasodilation only at more severe levels of arterial desaturation. Hypoxia is a complementary, physiologically distinct vasoactive stimulus. The use of hypoxic and inhaled-gas perturbations in MR imaging in fact predates fMRI itself. The original demonstrations of the BOLD effect by Ogawa and colleagues at 7 T in rodent brain used anoxia and inhaled-gas manipulations to establish that endogenous deoxyhemoglobin acts as a paramagnetic susceptibility contrast agent visible in vivo, defining the contrast mechanism that all subsequent BOLD imaging relies on (Ogawa, Lee, Nayak, et al., 1990; Ogawa, Lee, Kay, et al., 1990). Turner and colleagues extended this to time-resolved gradient-echo EPI in cat brain at 4 T, showing that BOLD could resolve oxygenation dynamics on the respiratory time scale (Turner et al., 1991), and Stehling and colleagues transferred the paradigm to humans using voluntary apnea, reporting 13–20% EPI signal attenuation as arterial deoxyhemoglobin rose (Stehling et al., 1993). Two mid-1990s animal studies grounded the BOLD–saturation relationship quantitatively: Jezzard et al. validated 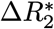 against direct optical SaO_2_ measurement through a cranial window in cat brain across anoxia, apnea, and hypercapnia, identifying cerebral blood volume (CBV) as a distinct contributor to the BOLD response to hypoxia (Jezzard et al., 1994); Prielmeier et al. mapped the 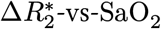 relationship in rat brain and showed that it is approximately linear over the mild-to-moderate hypoxic range and saturates under severe sustained deoxygenation (SaO_2_<=75% over several minutes), where compensatory CBF responses limit local accumulation of deoxyhemoglobin (Prielmeier et al., 1994). The first quantitative human characterization of fixed-inspired hypoxic BOLD followed shortly thereafter, with Rostrup and colleagues reporting robust 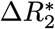 responses across cortex, deep gray matter, white matter, and the sagittal sinus at FiO_2_ = 10% alongside a 40% increase in total CBF,quantified by phase-contrast MRI at the bilateral internal carotid and vertebral arteries, 1 cm above the carotid bifurcation, at FiO_2_ = 10%; each measurement was preceded by a 2-min equilibration period and integrated over a 5–6 min phase-contrast scan (Rostrup et al., 1995). Using this same method, they observed minimal changes in CBF at delivery concentrations of 16%, 20%, 50%, and 100%. While this work established early evidence that hypoxia induced changes in CBF depends on the concentration of delivered O2 across a range of fixed-inspired oxygen levels, the changes they observed were at sustained exposures over roughly 8 minutes. Thus, the onset timing of this CBF response remained unresolved by Rostrup and colleagues. Almost two decades later, Harris and colleagues used pulsed arterial spin labeling to characterize the temporal dynamics of the CBF response to a 20-min hypoxic step (dynamic end-tidal forcing to PETO_2_ = 50 mmHg, achieving mean SpO_2_ ≈ 83%) and found that the CBF response was substantially delayed relative to the BOLD signal, with an onset delay of approximately 3 minutes for CBF versus 24 seconds for 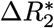 (Harris et al., 2013).

### 1.5 Hypoxia as a complementary vasoactive stimulus

Both hypoxic and hypercapnic BOLD resonses are driven by a combination of changes in deoxyhemoglobin concentration and CBF. However, the hypoxic BOLD response has been established as mechanistically distinct from the hypercapnic BOLD response. The hypercapnic BOLD response is driven primarily by CBF induced washout of deoxyhemoglobin, a result of vasodilation via CO_2_ mediated increases in blood acidity. In contrast, the hypoxic BOLD response has been argued to primarily reflect deoxyhemoglobin accumulation, accompanied by changes in CBF depending on both the dose administered (Rostrup et al., 1995) and the duration of administration (Harris et al., 2013). At very mild levels — for example, FiO_2_ ≈ 16%, where arterial desaturation is small (SaO_2_ ≥ 90%) — the CBF response has been argued to be modest enough that the BOLD signal change can be reasonably attributed to tissue deoxyhemoglobin alone, an assumption exploited to map relative cerebral blood volume (Wise et al., 2010). The temporal dynamics of hypoxia-induced CBF changes have also been characterized by Harris and colleagues using pulsed arterial spin labeling during a 5-min normoxic baseline followed by a continuous 20-min hypoxic step (dynamic end-tidal forcing to PETO_2_ = 50 mmHg, achieving mean SpO_2_ ≈ 83%) and an 8-min recovery, who found that the CBF response to hypoxia is substantially delayed relative to the BOLD signal — with an onset delay of approximately 3 minutes for CBF versus 24 seconds for 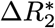 — implying that the early portion of any hypoxic BOLD response is likely dominated by deoxyhemoglobin rather than by flow (Harris et al., 2013). More recently, under the assumption that substantial CBF changes in response to hypoxia occur after a delay of around 180 seconds (Harris et al., 2013), a shorter transient step of moderate hypoxia (SaO_2_ ~= 80%) lasting 60 seconds has been used as a deoxyhemoglobin “contrast bolus” for arterial arrival-time mapping (Bhogal et al., 2022; Duffin et al., 2024; Sayin et al., 2023; Sayin et al., 2025; Stumpo et al., 2024). This method of hypoxia-induced deoxyhemoglobin perfusion has been argued to show strong agreement with “gold-standard” gadolinium-based dynamic susceptibility contrast methods in the same subjects (Sayin et al., 2023; Stumpo et al., 2024).

Across this body of work, the BOLD signal itself has been modeled in distinct ways. Bhogal and colleagues regressed the BOLD time series directly on the measured PETO_2_ trace to produce voxelwise CVR-style maps in units of %ΔBOLD per mmHg PETO_2_ (Bhogal et al., 2022). The remaining studies routed the hypoxic step into a dynamic susceptibility contrast (DSC)-style perfusion-modeling pipeline rather than a BOLD-CVR regression: Sayin and colleagues applied a Hill transformation to the PETO_2_ trace to derive an arterial SaO_2_ / [dOHb] time series and then used the **derived** trace itself as a model arterial input function (AIF) for VERBENA perfusion deconvolution (Sayin et al., 2023) — an approach later extended to compute perfusion metrics (rCBV, rCBF, MTT) directly from a step-response fit (Duffin et al., 2024) and to classify hemodynamic failure in steno-occlusive disease (Sayin et al., 2025). Stumpo and colleagues differed in AIF derivation, instead selecting a vascular voxel over the middle cerebral artery as the AIF and using a linear BOLD-on-PETO_2_ regression only to extract the percent-signal-change contrast time-course feeding their DSC pipeline (Stumpo et al., 2024). The directly measured pulse-oximetry SpO_2_ trace, by contrast, has not been used as a BOLD regressor in any of these studies.

### 1.6 Current gaps in the literature

Taken together, these findings identify several open questions about the hypoxic BOLD response: notably, the degree to which the CBF response to hypoxia impacts BOLD signal, and how the deoxyhemoglobin and CBV/CBF contributions to BOLD can be disentangled without an independent spin-echo or ASL measurement. Some work has assumed minimal to no CBF response to their hypoxic manipulations on the grounds of delivering a mild dose of hypoxia (e.g., 16% O_2_) (Wise et al., 2010) or on the grounds that a brief 1 minute hypoxic bolus is shorter than the previously observed 3 minute CBF response onset observed by Harris and colleagues (Bhogal et al., 2022; Duffin et al., 2024; Harris et al., 2013; Sayin et al., 2023). However, the underlying physiological factors contributing to the hypoxic BOLD response is still being characterized, and the body of work up until now has shown it varies widely among individuals. Given this uncertainty, we choose to refer to the hypoxic BOLD response in our present study as hypoxic CVR rather than assume it purely reflects intravascular deoxyhemoglobin content.

Additionally, a comparison of modeling approaches typically implemented in the literature to characterize the hypoxic BOLD response is warranted. While the PETO_2_ trace has been used by some (Bhogal et al., 2022), others have used Hill-style transformations to derive an estimate of arterial O_2_ saturation (Balaban et al., 2013; Sayin et al., 2023). The relationship between PETO_2_ and BOLD signal may not necessarily be as direct as PETCO_2_, thus the choice of regressor in modeling the hypoxic BOLD response deserves careful consideration. To our knowledge, no published study has compared regressor choices within a single dataset, and a Severinghaus-derived SaO_2_ trace has not yet been used as a primary BOLD GLM regressor for hypoxic CVR mapping. The Severinghaus 1979 formulation is standardly applied together with a full three-term Bohr correction for PCO_2_, pH, and temperature, whereas the Hill-equation fits used in the perfusion-imaging lineage (Balaban et al., 2013) handle only pH; the two formulations agree closely at fixed baseline conditions but diverge when PCO_2_ is permitted to vary freely, as it is under fixed-inspired gas delivery. We motivate this choice in more detail in §4.2.

A second, related gap concerns reproducibility — particularly at ultra-high field. The increased BOLD sensitivity and finer attainable spatial resolution at 7 T enable mapping of CVR with regional specificity, including in challenging regions such as the brainstem and white matter (Thomas et al., 2014). However, these gains come with greater *B*_0_ and *B*_1_ inhomogeneity artifacts that can degrade image quality, particularly in regions distant from the receive coil array such as the brainstem (Zuo et al., 2013). A handful of studies have used the field strength to characterize the hypercapnic BOLD response in detail — notably Bhogal and colleagues, who mapped voxelwise hypercapnic CVR amplitude and lag to a hypercapnic block paradigm at 7 T (Bhogal et al., 2015). Test–retest reproducibility of gas-challenge BOLD CVR at 7 T, however, remains largely uncharacterized even for hypercapnia alone, and the reproducibility of hypoxic CVR at 7 T has not been reported. To our knowledge no published human study has combined hypercapnic and hypoxic gas challenges in a single 7 T BOLD acquisition for voxelwise CVR mapping, let alone characterized the cross-session reproducibility of such maps. This is an important question given the potential clinical applications for hypoxic CVR mapping. Hypoxia-sensitive vascular measures are of particular interest in cohorts where cerebral oxygen extraction may be altered — for example, in patients with post-COVID conditions, where elevated cerebral oxygen extraction fraction has recently been reported and linked to reduced locomotor performance (Liu et al., 2024). The present reproducibility characterization is intended as a methodological foundation for such applications, including an ongoing large-cohort long-COVID study in our group that combines hypercapnic and hypoxic gas challenges to probe both dimensions of cerebrovascular reactivity in this population.

### 1.7 The present study

In the present work, we present and characterize such a combined paradigm: a continuous-flow, fixed-inspired gas manipulation protocol that delivers alternating hypercapnic and hypoxic gas boluses, separated by normocapnic/normoxic medical-air recovery periods, in a single 18-minute multi-echo, multiband BOLD acquisition at 7 T, with breath-by-breath end-tidal CO_2_/O_2_ and SpO_2_ monitoring. The multi-echo acquisition is important at 7 T because it enables optimal echo combination for improved signal-to-noise ratio and recovered coverage in high-susceptibility regions where a single-echo scan would lose signal to dropout, and because it produces a voxelwise 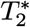 map that we use for objective signal-quality-based mask refinement (§2.6) and a per-volume 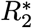 time series that provides an internal cross-check on the vascular origin of the observed BOLD excursions (§3.2). We describe the resulting voxelwise hypercapnic (CVR_HC_) and hypoxic (CVR_HO_) CVR maps under three physiologically motivated trace-based GLMs that differ in the regressor used to express the inhaled gas stimulus: a canonical end-tidal-trace (PETCO_2_ / PETO_2_) GLM, a directly measured pulse-oximetry SpO_2_ GLM, and a Severinghaus-derived SaO_2_ GLM that incorporates the in-vivo oxygen dissociation curve with Bohr correction. In parallel to the trace-based models, we additionally estimate voxelwise %ΔBOLD (and, when normalized to the delivered change in end-tidal gas concentration, CVR) using a finite impulse response (FIR) GLM. Unlike the trace-based models, the FIR approach does not use the recorded end-tidal or peripheral-saturation traces as regressors — it estimates the vascular response directly from the BOLD signal around each bolus event — and therefore avoids two common measurement-related sources of error in gas-challenge CVR: variable end-tidal trace quality (mask leaks, sampling delays) and the temporal alignment of each trace to the BOLD signal. Reproducibility of ROI-wise CVR estimates across two imaging sessions is then assessed for both conditions using intraclass correlation, with the goal of establishing the combined paradigm as a feasible and reliable tool for probing two complementary axes of cerebrovascular reactivity in health and disease.

## Methods

### 2.1 Participants

Thirteen healthy adult volunteers were initially enrolled in the reproducibility cohort, recruited from the greater Montreal area via general Facebook advertisements. Eligibility criteria included age ≥ 18 years; no history of cardiovascular, cerebrovascular, or severe respiratory disease; no severe obesity (operationally taken as BMI > 35); no clinical contraindications to the gas-challenge paradigm; no MRI contraindications; and the ability to wear and tolerate the breathing mask for the duration of the scan. Participants were additionally required not to be taking beta-blockers, which can directly affect autonomic function. Two participants were excluded from the analyzed cohort due to poor end-tidal forcing and/or mask fit during the gas-challenge paradigm, which produced unreliable gas-trace sampling, leaving *n* = 11 in the final analyzed sample. All participants gave written informed consent under a protocol approved by the institutional research ethics board.

The analyzed sample (*n* = 11) was 40.9 ± 7.2 years old (range 31–57), seven female and four male (sex assigned at birth), and predominantly right-handed (ten right, one left). Self-identified ethnicity was Caucasian in nine participants, South Asian in one, and Middle Eastern / North African in one. Highest education was Graduate degree for six participants, Undergraduate degree for four, and Secondary school for one; occupation was worker for eight, student for two, and unemployed for one. Ten participants were non-smokers and one was a former smoker, and none had a history of cardiovascular or cerebrovascular disease or other chronic conditions known to confound cerebrovascular reactivity. Cognitive screening was performed remotely over video call using the MoCA-Blind, a 22-point version of the Montreal Cognitive Assessment that excludes items requiring visual stimuli (trail-making, cube copy, clock drawing, and naming). The cohort mean was 20.0 ± 1.7 (range 18–22); all participants met or exceeded the MoCA-Blind cutoff of ≥ 18 for cognitively normal performance (equivalent to the standard MoCA cutoff of ≥ 26 on the 30-point version) (Wittich et al., 2010). BMI, height, and weight were not recorded uniformly across the cohort, but all participants were screened at enrolment to meet the inclusion limits described above.

### 2.2 Data Acquisition

Participants were scanned across two separate sessions (1 week to 3 months apart) on a 7 T Siemens MAGNETOM Terra scanner at the Montreal Neurological Institute Brain Imaging Center in Montreal, Quebec. High-resolution anatomical images were acquired using a T1-weighted MP2RAGE sequence (voxel size 0.7 mm3, TR/TE = 6000/2.73 ms, in-plane acceleration factor R = 3, field of view 240 × 240 × 168 mm, matrix 342 × 342 × 240). Whole-brain BOLD fMRI images were acquired using a multi-GRE, multiband sequence (TR = 1.72 s, TE = 11, 27, and 44 ms, isotropic voxel size 1.9 mm, multiband acceleration factor = 3, in-plane acceleration factor R = 3, flip angle 67°, field of view 224 × 224 mm, echo train length 29).

These structural and functional MRI sequences were acquired with concurrent end-tidal CO_2_/O_2_ monitoring using a GEMINI gas analyzer (CWE Inc.). Participants wore a facemask to allow control of inspired and expired gas concentrations. Physiological monitoring was synchronized to SpO_2_ data sampled with a Nonin medical-grade pulse oximeter using an MRI-compatible fiberoptic finger sensor. End-tidal gas and SpO_2_ data were synchronized with scanner TTL pulses using a BIOPAC MP160 physiological acquisition unit to align physiological recordings with fMRI volumes for subsequent analyses.

### 2.3 Fixed-Inspired Gas Manipulation

Inspired gas was delivered at 25 L/min using a custom gas mixer device built in our lab. For most of the scan, participants breathed medical air through the fixed facemask. During the gas manipulation scan, which lasted 18 minutes, participants received two boluses of a hypercapnic gas mixture (5% CO_2_, 21% O_2_, balance N_2_) and two boluses of a hypoxic gas mixture (10% O_2_, balance N_2_).

The hypercapnia boluses lasted 2 minutes and were followed by a 1-minute recovery period, whereas the hypoxia boluses lasted 3 minutes and were followed by a 3-minute recovery period. All scans began with a 1-minute medical-air baseline before four boluses were delivered in a fixed order: hypercapnia, hypoxia, hypercapnia, hypoxia.

### 2.4 Preprocessing

All structural and functional preprocessing was performed with *fMRIPrep* 25.1.4 (Esteban et al., 2018; Esteban et al., 2019) (built on *Nipype* 1.10.0 (Gorgolewski et al., 2011; Gorgolewski et al., 2018) and *Nilearn* 0.11.1 (Abraham et al., 2014)). The following paragraphs describe the operations relevant to the CVR analysis; full workflow details are documented in the *fMRIPrep* documentation.

#### 2.4.1 *B*_0_ Inhomogeneity Mapping

For each session, a *B*_0_-nonuniformity map (fieldmap) was derived from the two spin-echo EPI references acquired with opposing phase-encoding polarity using `topup` (Andersson et al., 2003). This fieldmap was subsequently used for susceptibility distortion correction of the multi-echo BOLD runs.

#### 2.4.2 Anatomical Data Preprocessing

For each subject, the T1-weighted MP2RAGE image acquired at each session was corrected for intensity non-uniformity with `N4BiasFieldCorrection` (Tustison et al., 2010) (ANTs 2.6.2 (Avants et al., 2008)), skull-stripped using the `antsBrainExtraction.sh` workflow with MNI152NLin2009cAsym as the target template, and segmented into cerebrospinal fluid, white matter, and gray matter using `fast` (Zhang et al., 2001). A subject-level anatomical reference was computed by robust registration of the two per-session T1w images with `mri_robust_template` (Reuter et al., 2010). Cortical surfaces were reconstructed with `recon-all` (Dale et al., 1999), refined against a FLAIR image for improved pial-surface accuracy, and further refined via a Mindboggle procedure (Klein et al., 2017). Volumetric spatial normalization to MNI152NLin2009cAsym (accessed via *TemplateFlow* 24.2.2 (Ciric et al., 2022; Fonov et al., 2009)) was performed by nonlinear registration with `antsRegistration`.

#### 2.4.3 Functional Data Preprocessing

For each BOLD run, a reference volume was generated from the shortest echo, and head-motion parameters were estimated with `mcflirt`(Jenkinson & Smith, 2001). The subject-specific fieldmap was rigidly aligned to the BOLD reference to enable susceptibility distortion correction. The BOLD reference was then co-registered to the T1w reference using boundary-based registration (`bbregister`(Greve & Fischl, 2009)). Head-motion, susceptibility distortion, and coregistration transforms were composed and applied in a single cubic B-spline resampling step using `nitransforms`to produce BOLD data in MNI152NLin2009cAsym space.

After per-echo preprocessing, the three echoes were optimally combined across echoes using the 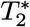-weighted combination implemented in *tedana* (DuPre et al., 2021), producing a single BOLD time-series per volume with improved signal-to-noise ratio and recovered signal in high-susceptibility regions (orbitofrontal cortex, temporal poles, ventral cerebellum, and brainstem) where a single-echo acquisition would lose signal to dropout. The voxelwise 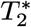 map produced during optimal combination was retained for the tissue-mask refinement described in §2.6. All GLMs (§2.5) were fit to this optimally combined BOLD time series. In parallel, a per-volume 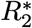 time series was estimated separately from the three preprocessed echoes for visualization of the hemodynamic response independently of the *S*_0_-modulated BOLD signal (§3.2); see Figure 2 caption for the estimation formulation.

### 2.5 CVR Estimation

CVR is typically defined as the change in BOLD signal per unit change in end-tidal CO_2_ (ETCO_2_). In the present study, we additionally quantified change in BOLD signal per unit change in three candidate hypoxia regressors: end-tidal O_2_ (ETO_2_), peripheral oxygen saturation (SpO_2_), and a Severinghaus-derived arterial oxygen saturation (SaO_2_) trace. To distinguish these measures, we refer to CO_2_-based cerebrovascular reactivity as CVR_HC_ and O_2_-based reactivity as CVR_HO_, reflecting BOLD responses to hypercapnia and hypoxia, respectively. We fit three trace-based GLMs that share the same design-matrix structure but differ in the hypoxia regressor (§2.5.1 and §2.5.2), and a finite impulse response (FIR) GLM that makes no assumption about response shape and does not rely on the recorded physiological traces, serving both as a trace-free estimator of %ΔBOLD and as a reference for best-possible model fit (§2.5.3).

#### 2.5.1 Canonical End-tidal Trace General Linear Model Analysis

CVR is commonly analyzed using a GLM similarly to task-evoked fMRI, except that the ETCO_2_ trace is included as a regressor in the design matrix. This approach requires temporal alignment of the end-tidal traces to the BOLD signal to account for lung-to-brain transit time. As is standard practice in the BOLD CVR literature (Liu et al., 2019), we performed this alignment by computing the cross-correlation between each end-tidal trace and a whole-brain gray-matter mean BOLD reference time-series across a range of lags, and selecting the lag that maximized the cross-correlation. Voxelwise lag optimization has been used in the literature to recover regional variation in lung-to-brain transit time (Bright & Murphy, 2013), but is not typically applied to voxel-scale hypercapnic CVR mapping because per-voxel lag estimates can be noisy in low-SNR regions and voxelwise optimization substantially increases the number of fitted parameters and computational cost (Liu et al., 2019). We therefore restricted alignment to a single subject-level lag per end-tidal trace, applied uniformly across voxels.

Applying this procedure across sessions and subjects, gray-matter BOLD lagged the ETCO_2_ regressor by a median of 9.0 s (mean 7.9 ± 4.8 s), consistent with the combined gas-analyzer sampling, lung-to-brain transit, and cerebrovascular response times reported in prior CVR work (Bright & Murphy, 2013; Liu et al., 2019). The Severinghaus-derived SaO_2_ regressor showed a comparable lag (median 6.5 s; mean 8.4 ± 9.7 s), as did ETO_2_ (median 6.0 s; mean 4.3 ± 9.1 s), although the O_2_ traces produced broader, less well-defined correlation peaks than the ETCO_2_ trace. The peripheral pulse-oximetry SpO_2_ regressor required a large positive shift (median +35.5 s; mean +41.4 ± 28.8 s) — matching the well-documented 30 s response delay of finger pulse oximetry relative to arterial saturation and reflecting peripheral measurement delay rather than a neurovascular process.

For the present analysis, both ETCO_2_ and ETO_2_ were aligned to the gray-matter reference signal separately before a single GLM was run with both traces as regressors. Both regressors were modeled simultaneously to estimate their independent contributions to the BOLD signal during the combined gas manipulation.

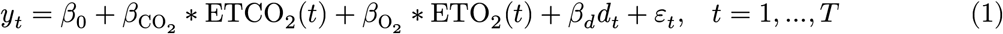

CVR_HC_ was defined as the percent signal change per mmHg change in ETCO_2_. Mirroring Liu and colleagues, the minimum of the ETCO_2_ trace — computed across the entire gas-manipulation scan, which for our fixed-inspired paradigm corresponds to the resting normocapnic (medical-air) baseline — was included in the denominator so that reactivity is referenced to the baseline physiological state rather than to ETCO_2_ = 0 (Liu et al., 2019). A similar definition was used for CVR_HO_, using the maximum of the ETO_2_ trace (again computed across the entire scan) as the baseline state under hypoxia (Liu et al., 2019).

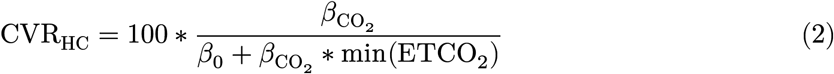

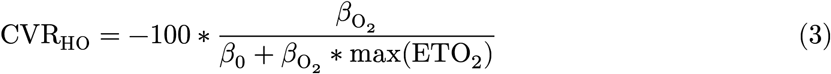

#### 2.5.2 SpO_2_ **and Severinghaus-derived** SaO_2_ **GLM Analyses**

The relationship between ETO_2_ and the hypoxic BOLD response is less direct than that between ETCO_2_ and the hypercapnic BOLD response, in part because the BOLD signal during hypoxia is mediated by changes in intravascular oxygen saturation rather than by alveolar partial pressure of O_2_ per se (Prielmeier et al., 1994). To address this we additionally fitted two trace-based GLMs that shared the same design-matrix structure as the canonical end-tidal trace GLM but replaced the ETO_2_ regressor with: (i) the measured peripheral oxygen saturation (SpO_2_) trace; and (ii) a Severinghaus-derived arterial oxygen saturation (SaO_2_) trace computed from the end-tidal traces themselves.

The SpO_2_ GLM used the pulse-oximeter SpO_2_ signal in place of ETO_2_:

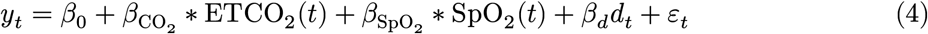

The SaO_2_ trace was derived from the end-tidal traces using the Severinghaus oxyhemoglobin dissociation equation (Severinghaus, 1979) with a Bohr correction that accounts for CO_2_-dependent shifts in the oxyhemoglobin dissociation curve. The standard Severinghaus form

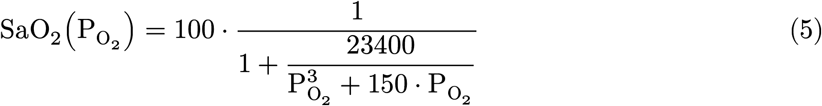

was applied to a Bohr-corrected virtual standard-condition PO_2_ rather than directly to the measured ETO_2_:

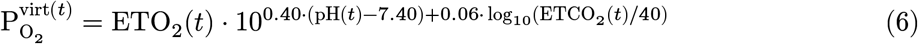

where the pH was approximated from the concurrent ETCO_2_ trace via the standard acute respiratory acid–base relationship pH(*t*) = 7.40 − 0.008 ⋅ (ETCO_2_(*t*) − 40). The 0.40 (pH) and 0.06 (PCO_2_) coefficients are those given by Severinghaus and capture the leftward shift of the oxyhemoglobin dissociation curve under acidotic conditions. The SaO_2_ GLM then used the derived SaO_2_ trace as the hypoxia regressor:

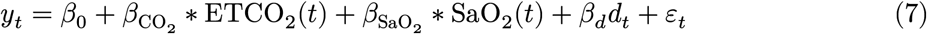

In both saturation-trace GLMs the ETCO_2_ regressor and the nuisance terms (linear drift, intercept) were retained as in the canonical end-tidal trace GLM. The SpO_2_ and SaO_2_ traces were each aligned to the gray-matter reference signal by the same cross-correlation procedure used for ETO_2_. CVR_HO_ for each model was computed analogously to the canonical end-tidal definition, replacing max(ETO_2_) in the denominator with the maximum (resting) value of the corresponding saturation trace:

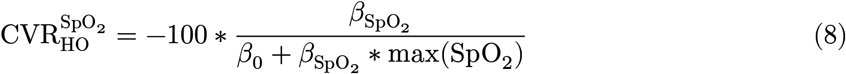

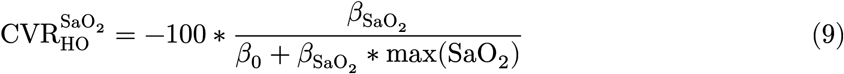

#### 2.5.3 Finite Impulse Response General Linear Model Analysis

In parallel to the three trace-based GLMs above, we fit a finite impulse response (FIR) GLM (Glover, 1999) that estimates the BOLD response to each gas bolus directly from the observed data, without using the recorded end-tidal or peripheral-saturation traces as regressors. Instead of one regressor per gas, the FIR model represents the vascular response as a sequence of time-lagged coefficients following each bolus onset (one *β* per time-lag per condition), from which the mean %ΔBOLD in each response window is extracted (Eqs. 12 and 13). This trace-free formulation avoids two measurement-related error sources present in trace-based fits — variable end-tidal trace quality (mask leaks, sampling delays) and the need to temporally align each trace to the BOLD signal — at the cost of not producing a CVR value normalized to the delivered gas dose unless the corresponding ΔETCO_2_ or ΔETO_2_ is applied post-hoc (§3.4). Because the FIR design matrix has substantially more free parameters than the trace-based GLMs (tens of *β*s per condition versus one *β* per gas regressor), it can also fit session-specific noise more freely, a trade-off returned to in the Results and Discussion (Hoge et al., 1999).

Under this framework, hypercapnia and hypoxia boluses were treated as discrete events, with each bolus onset assumed to initiate a transient vascular response that evolves over time. Separate binary event indicators were defined for each condition:

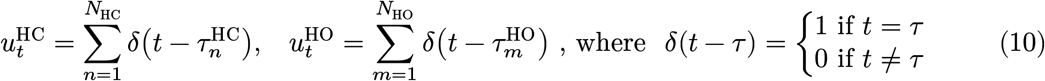

The BOLD signal can then be modeled as the linear superposition of time-lagged responses to these events:

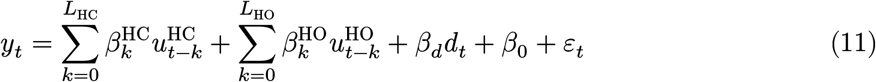

Here, 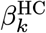 and 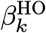 correspond to the estimated BOLD response at lag *k* following the onset of a hypercapnia or hypoxia bolus, respectively.

In the FIR framework, percent signal change for hypercapnia and hypoxia was estimated by averaging a subset of lag coefficients corresponding to a steady-state window of interest, dividing by the model intercept, and multiplying by 100 (see Eqs. 12 and 13). In the present work, the response window was taken as the second half of the bolus delivery period for each gas type.

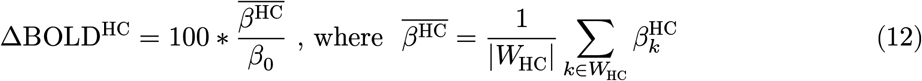

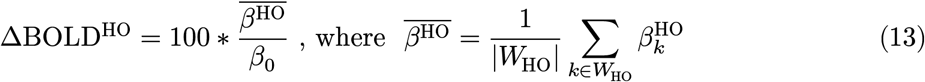

CVR_HC_ and CVR_HO_ were then estimated by dividing the percent signal change for each condition by the corresponding change in end-tidal CO_2_ or O_2_. Change in end-tidal gas was estimated by subtracting the mean value over the 30 seconds before bolus onset from the mean value over the selected response window.

### 2.6 Reproducibility Analysis

#### 2.6.1 ROI-wise ICC Analyses

Across-session reproducibility of voxelwise CVR maps was assessed descriptively by computing intraclass correlation coefficients (ICCs) per region of interest (ROI) across the two scanning sessions. For each (subject, session, ROI) the mean CVR was computed across in-mask voxels, with all CVR maps restricted to voxels whose mean 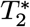 (from the tedana optimal-combination step, §2.4.3) fell inside the plausible-tissue window of 10 to 40 ms — a range that excludes signal-dropout zones (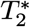 < 10 ms) and CSF / partial-volume edges (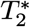 > 40 ms), producing a signal-quality-based mask objectively derived from the multi-echo acquisition rather than an anatomically defined atlas boundary.

ROIs were drawn from four atlases covering complementary spatial scales: the Harvard-Oxford cortical and subcortical atlases distributed with FSL (Desikan et al., 2006; Jenkinson et al., 2012; Makris et al., 2006), binarized at a probability threshold of 0.25 to reduce partial-volume effects; the FreeSurfer brainstem subdivisions (combined brainstem, pons, midbrain, superior cerebellar peduncle, and medulla); and the 58-nucleus Brainstem Navigator atlas (Hansen et al., 2024). ICC was additionally tabulated separately for gray-matter and white-matter tissue masks.

ICCs were computed in `pingouin`(Vallat, 2018) using the ICC(3, k) variant — a two-way mixed-effects model with averaged measurements (Shrout & Fleiss, 1979) — separately for hypercapnia and hypoxia, with the two scanning sessions treated as the raters. ICC values range from 0 to 1, with higher values indicating greater between-session consistency relative to between-subject variability; we adopt the conventions of 0.4, 0.6, and 0.75 as fair-, good-, and excellent-reproducibility thresholds for descriptive interpretation. The reported 95% confidence intervals are pingouin’s default parametric intervals derived from the *F*-distribution and are **not** corrected for multiple comparisons across the many ROIs per atlas. Accordingly, the resulting figures are interpreted descriptively — by the proportion of ROIs whose point estimate or lower 95% bound clears the 0.4 or 0.6 thresholds — rather than as formal hypothesis tests at each ROI. Statistical power for ICC interval estimation is necessarily limited by our sample size of 9–12 participants per (atlas, condition) pair, well below the *n* ≥ 30 commonly recommended for stable interval estimates (Koo & Li, 2016). The lower bound of *n* = 9 reflects incomplete pulse-oximetry data for two participants due to intermittent SpO_2_-device dropouts during the gas-manipulation scan; the SpO_2_-GLM and ΔSpO_2_ analyses were therefore restricted to the *n* = 9 subset with usable SpO_2_ recordings across both sessions, while the ET- and SaO_2_-GLM analyses used the full *n* = 11 sample.

## Results

### 3.1 Physiological Responses to Gas Manipulation

The fixed-inspired gas manipulation successfully induced robust and reproducible changes in end-tidal gas concentrations and peripheral oxygen saturation across participants and sessions (Figure 1, Table 1). Figure 1 shows group mean traces for ETCO_2_, ETO_2_, and SpO_2_ across the gas manipulation scan, overlaid with the inspired-gas concentrations delivered by the continuous-flow mixer. Both end-tidal traces show clear modulation in response to the gas boluses with consistent timing and magnitude across sessions, and the SpO_2_ trace shows a clear and consistent decrease across both hypoxia boluses.

**Table 1:** ICC summary for end-tidal gas measures across sessions.

| Measure | ICC | 95% CI | p-value | Mean | SD | N | Reps |
| --- | --- | --- | --- | --- | --- | --- | --- |
| $\Delta\text{ETCO}_2$ (Hypercapnia) | 0.71 | (0.26–0.91) | $4.43 \times 10^{-3}$ | 7.72 mmHg | 1.96 | 11 | 4 |
| $\Delta\text{ETO}_2$ (Hypoxia) | 0.57 | (–0.07–0.87) | $3.49 \times 10^{-2}$ | –72.42 mmHg | 7.08 | 11 | 4 |
| $\Delta\text{SpO}_2$ (Hypoxia) | 0.65 | (0.04–0.91) | $2.09 \times 10^{-2}$ | –12.5 % | 3.34 | 9 | 4 |
| $\Delta\text{SaO}_2$ (Hypoxia) | 0.95 | (0.87–0.98) | $1.68 \times 10^{-10}$ | –11.86 % | 5.45 | 11 | 4 |

**Figure 1:**
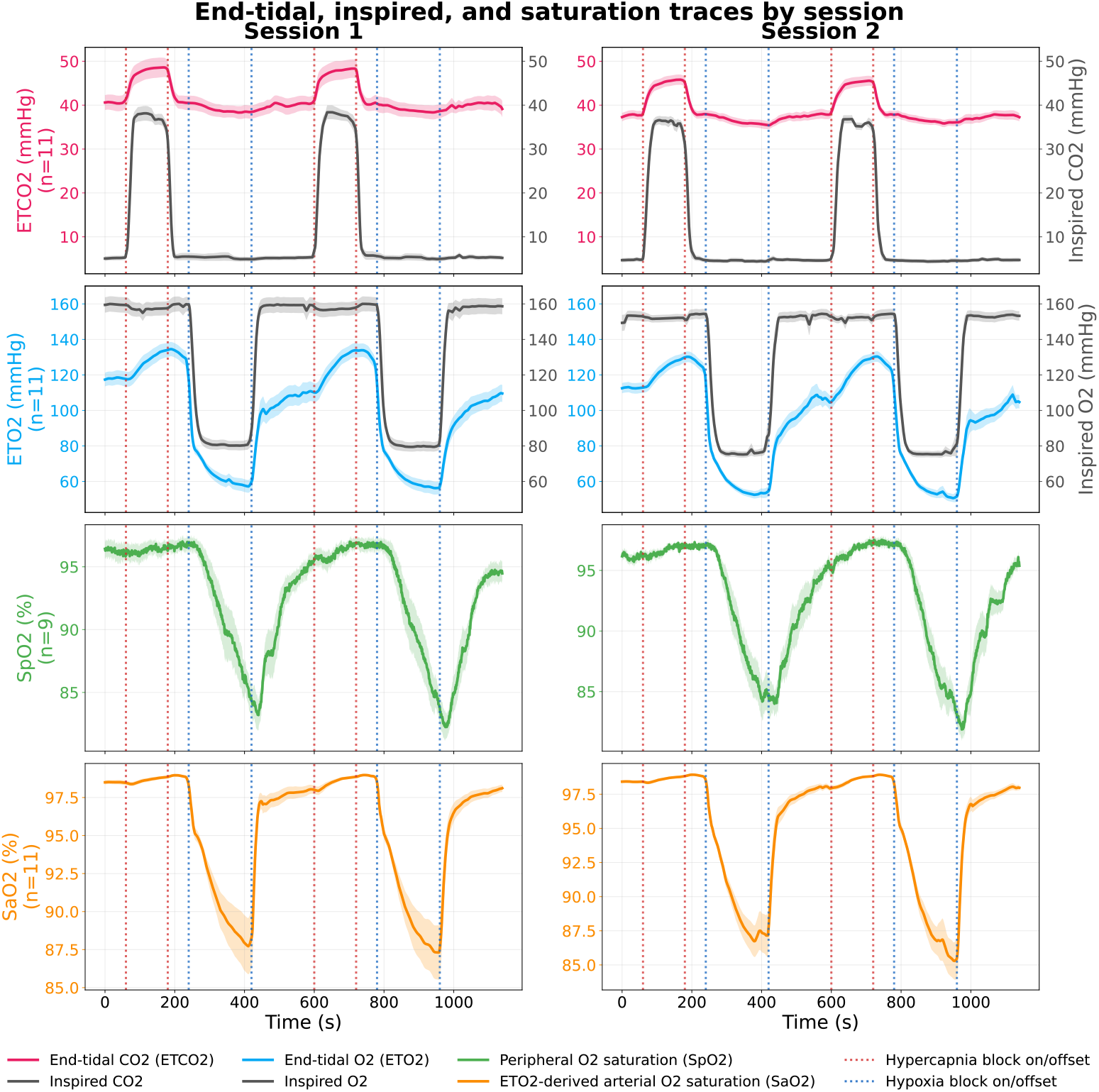
Group mean traces for End-Tidal CO_2_ (top), End-Tidal O_2_ (middle), and SpO_2_ (bottom), shown by session, overlaid with the corresponding inspired-gas concentrations of CO_2_ and O_2_ delivered by the continuous-flow mixer.

**Figure 2:**
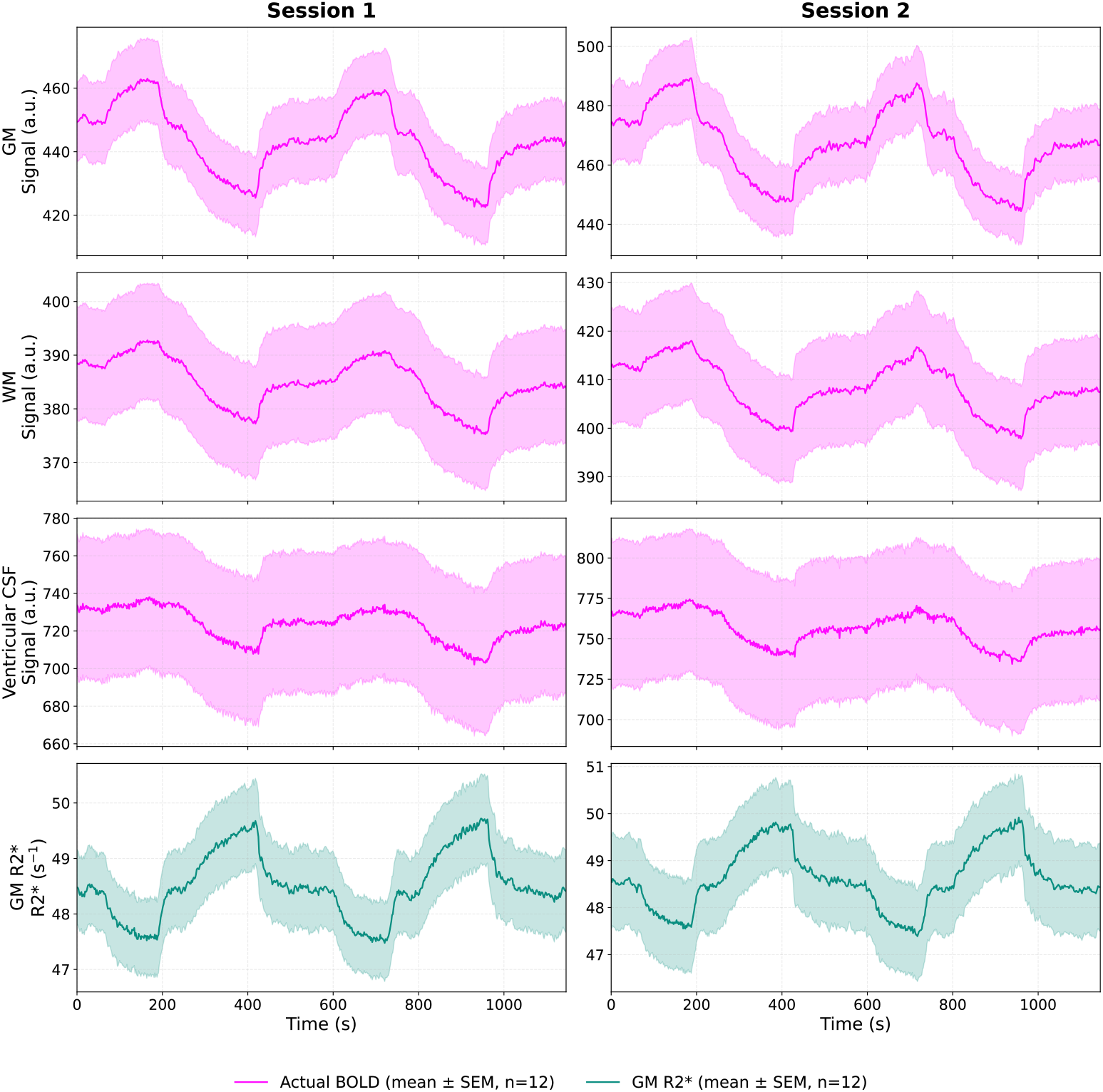
Group-mean tissue-signal time-series during the gas manipulation scan, for session 1 (left) and session 2 (right). Top three rows: mean BOLD signal within gray-matter (GM), white-matter (WM), and ventricular cerebrospinal-fluid (CSF) masks. Bottom row: mean gray-matter transverse relaxation rate (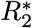, in s^−1^) estimated per volume by ordinary-least-squares log-linear fit across the three echoes and masked to the same GM tissue mask. Hypercapnia boluses produce positive BOLD excursions and hypoxia boluses produce negative BOLD excursions in GM and WM; the 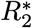 signal moves in the opposite direction to the GM BOLD signal, as expected for a signal whose sign is inverted relative to BOLD susceptibility contrast. WM responses are smaller in amplitude and temporally delayed relative to GM, but otherwise track the GM trace closely; response shape and magnitude are highly consistent across the two sessions.

To quantify the across-session reproducibility of the physiological response amplitudes themselves, we computed intraclass correlation coefficients (ICC(3, k); `pingouin`(Vallat, 2018)) across the four bolus repetitions per subject (two boluses per session, two sessions per subject) for the change in end-tidal CO_2_ during hypercapnia (ΔETCO_2_), the change in end-tidal O_2_ during hypoxia (ΔETO_2_), the change in peripheral pulse-oximetry saturation during hypoxia (ΔSpO_2_), and the change in the Severinghaus-derived arterial O_2_ saturation trace that drives the SaO_2_-GLM (ΔSaO_2_). All four amplitude measures showed positive across-session reproducibility (Table 1). The ΔETCO_2_, ΔETO_2_, and ΔSpO_2_ amplitudes fell in the fair-to-good range (ICCs 0.57 to 0.71). The Severinghaus-derived ΔSaO_2_ amplitude — the trace that actually drives the SaO_2_-GLM — was substantially more reproducible, falling in the excellent range (ICC = 0.95, 95% CI [0.87, 0.98]); the non-linear Severinghaus + Bohr transform appears to amplify subject-specific differences in baseline / nadir saturation while suppressing breath-by-breath envelope noise from the raw end-tidal trace, and we revisit this point in the Discussion.

### 3.2 BOLD Response to Gas Manipulation

The gas manipulation evoked clear, condition-specific BOLD responses in both gray matter (GM) and white matter (WM), and a mirror-image response in gray-matter transverse relaxation rate 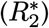, highly consistent across the two imaging sessions (Figure 2). Hypercapnia boluses produced positive BOLD excursions (and corresponding negative 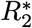 excursions) in both tissue compartments, while hypoxia boluses produced sustained negative BOLD excursions (and corresponding positive 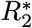 excursions) both responses returned toward baseline during the intervening normocapnic/normoxic recovery periods. Voxelwise 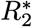 was estimated per volume by ordinary-least-squares log-linear fit of log 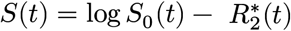 ⋅ TE across the three echoes (TE = 11, 27, and 44 ms), restricted to the union of GM, WM, and CSF tissue masks; the sign inversion between BOLD and 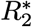 reflects their opposite dependence on tissue deoxyhemoglobin concentration and provides an internal cross-check on the vascular origin of the observed BOLD excursions.

Tissue compartments differed in the magnitude and timing of their responses in the manner expected from the underlying vascular and tissue properties. The GM signal showed larger response amplitudes and faster onsets and offsets for both hypercapnic and hypoxic boluses, consistent with its denser microvasculature and shorter transit times. The WM signal closely tracked the GM trace in shape and sign across all four boluses but with a smaller amplitude and a temporal lag, in line with prior reports of blunted, delayed CVR responses in WM relative to GM (Thomas et al., 2014). Comparison of the two sessions (left and right panels of Figure 2) shows that the magnitude, timing, and tissue-specific differences of the BOLD response — and of the 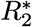 response — were reproduced across the repeat scan, with no systematic shift between sessions for either tissue type.

### 3.3 GLM Models Capture BOLD Responses to Gas Manipulation

All GLM models captured robust BOLD responses to the gas manipulation, with good model fit quality across participants and sessions. Figure 3 shows modeled vs actual BOLD time-series in gray matter for a representative participant (left panel) and for the group average across both sessions (right panel). As expected, the FIR GLM (top row) fit the shape of the BOLD response very well, including the high frequency content. The ET-GLM (second row) fit the shape of the BOLD response to hypercapnia fairly well, however it generally underestimated the BOLD response to hypoxia. The SpO_2_ (third row) and SaO_2_ (fourth row) GLMs showed higher fidelity in capturing the observed BOLD response to hypoxia than the canonical ET-GLM.

**Figure 3:**
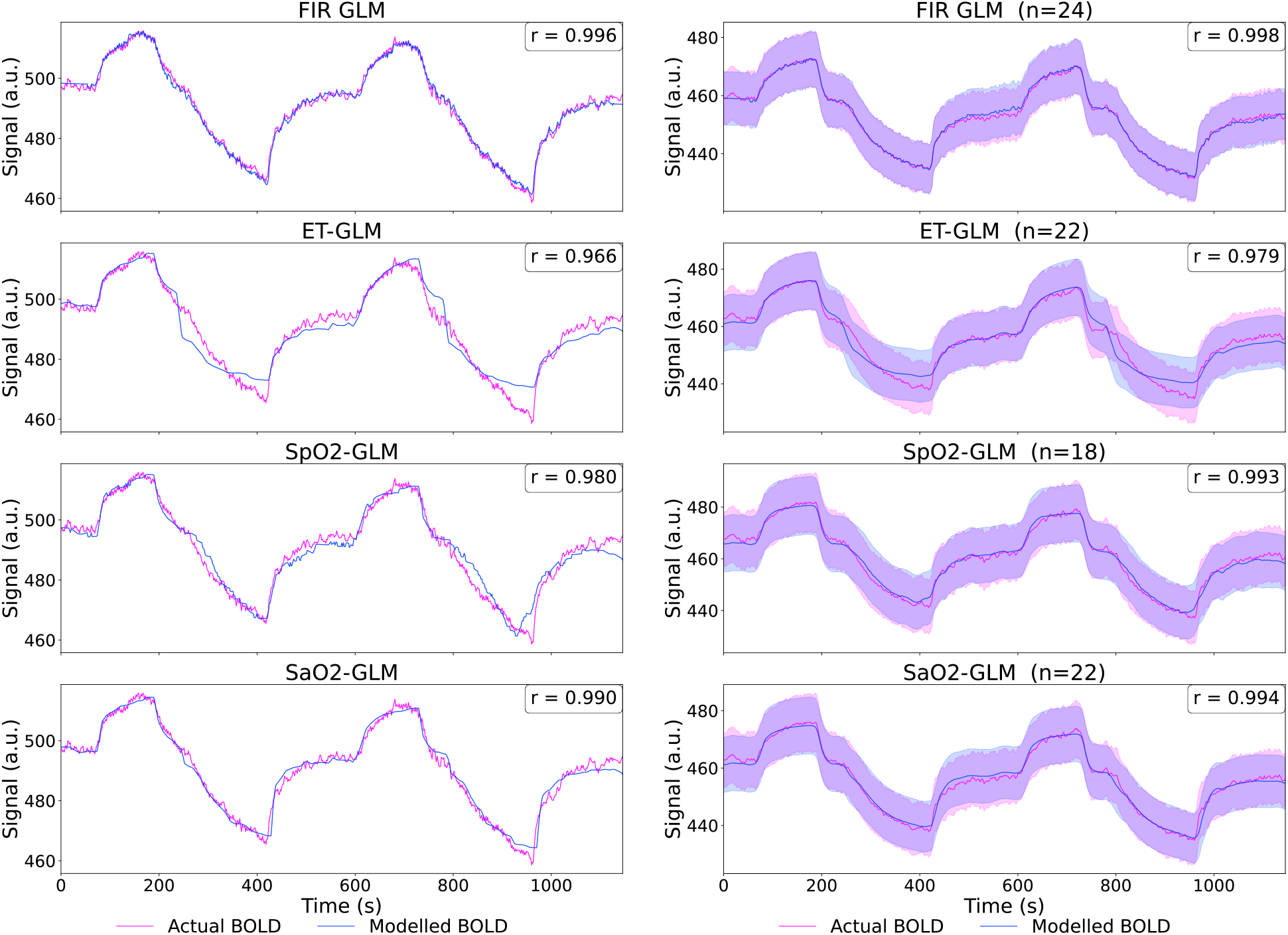
Modelled vs actual BOLD time-series in gray matter for the FIR GLM (top row), ET-GLM (second row), SpO_2_ GLM (third row), and SaO_2_ GLM (bottom row). Left column: representative participant. Right column: mean across participants and sessions.

To quantitatively compare model fit across the four GLMs we computed Pearson’s *r* between predicted and observed BOLD time-series in GM, both within-session (predicting the BOLD signal of the session on which the GLM was fit) and across sessions via two-fold cross-session validation (fitting on session A and predicting session B, and vice versa, with the two folds averaged in Fisher-*z* space). Pearson *r* was used as the primary metric because cross-session predictions inherit each session’s per-session *β*-weight scale and are therefore not directly comparable in raw *R*^2^ units; centered *R*^2^ is reported as a secondary measure in the Supplemental Material. Pairwise model contrasts were assessed with per-subject sign-flip permutation tests (10,000 permutations).

The FIR GLM produced the highest within-session *r* against observed GM BOLD (mean *r* = 0.976), reflecting its much larger degrees of freedom (one *β* per time-point), and significantly outperformed every other model under sign-flip permutation testing: FIR vs ET-GLM *p* = 0.001, FIR vs SaO_2_-GLM *p* = 0.002, and FIR vs SpO_2_-GLM *p* = 0.010 (Supplemental Figure S1). Among the trace-based models, the SaO_2_- and SpO_2_-driven GLMs (mean *r* = 0.960 each) both fit observed BOLD significantly better than the ET-GLM (mean *r* = 0.943; permutation *p* = 0.001 and *p* = 0.008 respectively).

Cross-session validation showed a distinct pattern (Supplemental Figure S2). All four models achieved high mean cross-session *r* (SaO_2_ = 0.939, SpO_2_ = 0.938, FIR = 0.935, ET-GLM = 0.925), with the SaO_2_ GLM edging out the others: under sign-flip permutation testing, SaO_2_ was the only model with pairwise advantages that survived, generalizing significantly better than both the ET-GLM (permutation *p* = 0.001) and the SpO_2_-GLM (*p* = 0.034). The FIR GLM’s within-session advantage did not carry over: all three FIR pairwise contrasts failed permutation testing (permutation *p* ∈ [0.05, 0.55]), placing FIR’s cross-session generalization statistically indistinguishable from the three trace-based models. This pattern — high within-session fit but no cross-session advantage — is consistent with the FIR model being more susceptible to session-specific noise than the trace-based models, an expected consequence of its higher degree-of-freedom design matrix. The trace-based models constrain the vascular response to follow the shape of the driving physiological trace, which effectively acts as an implicit temporal regularizer favoring cross-session generalization.

This pattern was evident at the individual-voxel level as well. We computed the same cross-session Pearson *r* per voxel for both the SaO_2_ and FIR GLMs and contrasted them with a paired t-test across subjects, false discovery rate (FDR)-corrected at *q* < 0.05 (Figure 4). In 58.0% of GM voxels (*n* = 120, 493 of 207, 863), the FIR GLM’s cross-session prediction was significantly less accurate than the SaO_2_ GLM’s; the FIR GLM was significantly more accurate than SaO_2_ in only 0.04% (*n* = 78); the remaining 42.0% showed no significant difference. The pattern was broadly distributed across the gray-matter mask rather than concentrated in a subset of regions, consistent with a general — rather than region-specific — tendency of the higher-DoF FIR fit to absorb session-specific noise.

**Figure 4:**
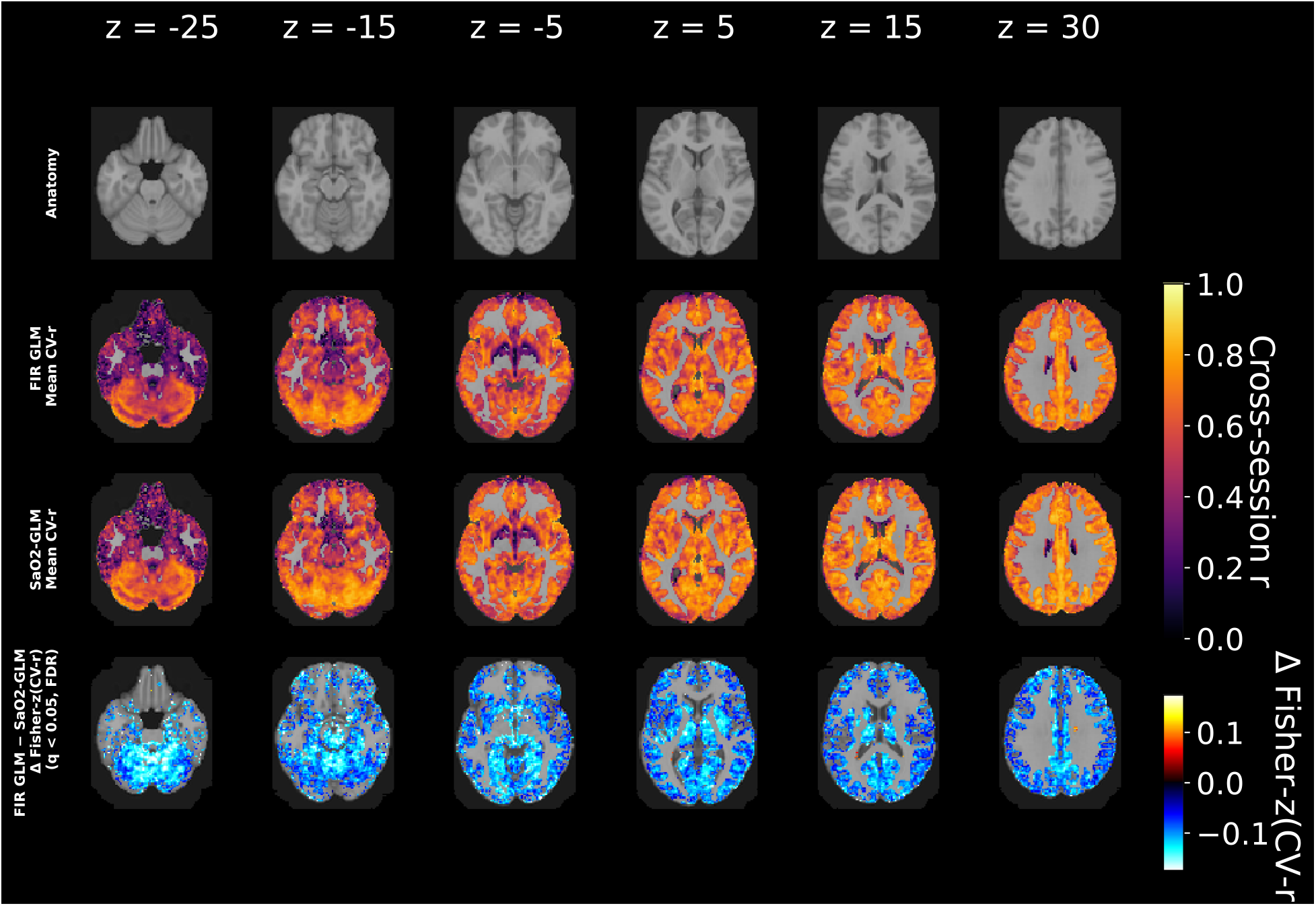
Voxelwise paired contrast of cross-session Pearson *r* between the FIR GLM and the Severinghaus-derived SaO_2_ GLM in gray-matter voxels, FDR-corrected at *q* < 0.05 across in-mask voxels. Negative values (blue) indicate voxels where the FIR GLM was less accurate than the SaO_2_ GLM at predicting cross-session BOLD; positive values (red) indicate the opposite. The blue distribution spans 58% of gray-matter voxels and is broadly distributed across the mask (red: 0.04%), consistent with the higher-degree-of-freedom FIR fit being more susceptible than the trace-based SaO_2_ GLM to session-specific noise.

### 3.4 Voxelwise CVR Maps

Voxelwise CVR maps were derived per subject and session from each of the four GLMs (FIR, ET-trace, SpO_2_, SaO_2_), and then averaged across subjects to produce group-mean maps for each session and each gas condition. The group-mean maps derived from the Severinghaus-derived SaO_2_ GLM are shown in Figure 5, since this model produced the most reproducible and best-generalizing predictions of the gas-induced BOLD response (§3.3). Group-mean maps from the FIR, ET-trace, and SpO_2_ GLMs are provided for comparison in Supplemental Figures S4, S5, and S6, respectively.

**Figure 5:**
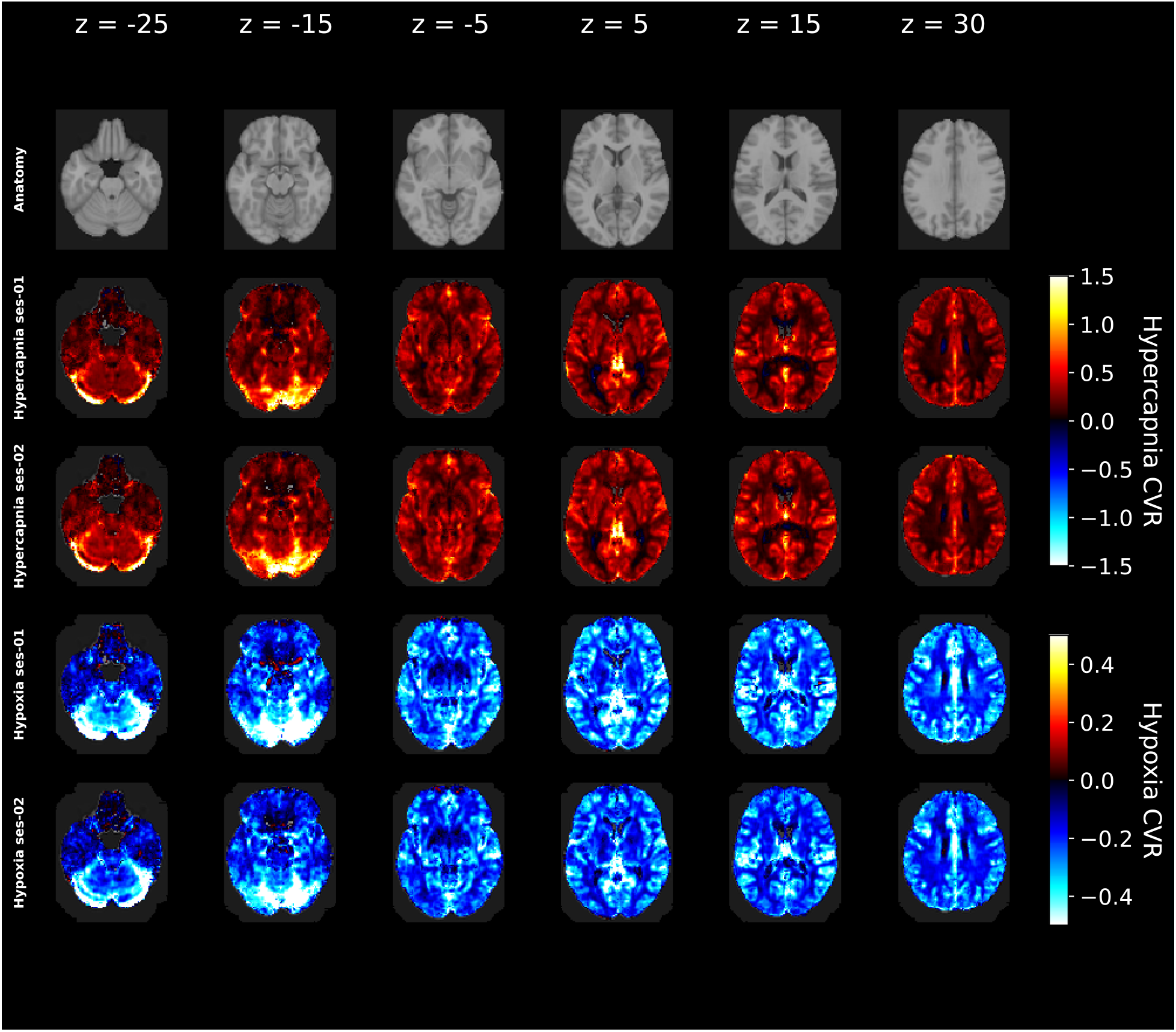
SaO_2_-GLM-derived group-mean voxelwise CVR maps for hypercapnia first session (top row), hypercapnia second session (second row), hypoxia first session (third row), and hypoxia second session (bottom row). CVR_HC_ is expressed in % ΔBOLD per mmHg ΔETCO_2_; CVR_HO_ is expressed in % ΔBOLD per Δ% SaO_2_. Together, these maps illustrate the expected positive gray-matter CVR_HC_ response and negative CVR_HO_ response reproduced across the two imaging sessions.

Voxelwise CVR_HC_ and CVR_HO_ were computed per Eqs. 2–9 for the trace-based GLMs and per Eqs. 12–13 for the FIR GLM (see Methods §2.5). Because the three hypoxia regressors carry different physical units —the ET-trace maps are expressed in % ΔBOLD per mmHg ΔETO_2_, whereas the SpO_2_ and SaO_2_ maps are expressed in % ΔBOLD per Δ% O_2_ saturation — the hypoxia CVR maps from different regressors are not directly numerically comparable but can be compared on the basis of spatial pattern and sign.

The SaO_2_-GLM group-mean maps (Figure 5) show robust positive CVR_HC_ in gray matter during hypercapnia (top two rows) and robust negative CVR_HO_ in gray matter during hypoxia (bottom two rows), with spatial patterns consistent across sessions. The FIR, ET-trace, and SpO_2_ GLMs yielded qualitatively similar voxelwise CVR maps to the SaO_2_ GLM, with the same gray-matter spatial coverage and the same session-to-session consistency for both conditions (Supplemental Figures S4–S6). The four GLMs’ CVR_HC_ maps in particular are visually near-identical, as expected: all three trace-based GLMs share the same ETCO_2_ regressor for hypercapnia and differ only in the choice of hypoxia regressor, so differences across models manifest almost exclusively in the CVR_HO_ maps.

### 3.5 Reproducibility Metrics

To assess across-session reproducibility of the CVR estimates produced by the Severinghaus-derived SaO_2_ GLM — the most generalizable predictor of the gas-induced BOLD response in our model-comparison analysis above — we computed intraclass correlation coefficients per ROI across the two scanning sessions. The analysis was restricted to the SaO_2_ GLM because the cross-session model-fit comparison (Supplemental Figure S2) identified it as the only model with a permutation-defensible cross-session generalization advantage at the gray-matter level, and as more accurate than the FIR GLM in 58% of GM voxels (Figure 4).

Across the four atlases the Harvard-Oxford cortical and subcortical ROIs were the most reproducible across sessions (median ICC = 0.71; 71% of cortical and subcortical (ROI × condition) tests with ICC > 0.6), followed by the FreeSurfer brainstem subdivisions and tissue means (median ICC = 0.63), with the smaller Brainstem Navigator nuclei the least reproducible (median ICC = 0.49; 37% with ICC > 0.6) — consistent with smaller voxel counts per ROI and reduced BOLD SNR in the brainstem. Across all 288 (ROI × condition) tests, 74% yielded ICC ≥ 0.4 (fair or better) and 30% yielded ICC ≥ 0.75 (excellent). Figure 6 summarizes ROI-wise ICC across the four atlases: (a) tissue compartments (GM, WM), (b) Harvard–Oxford cortical ROIs, (c) Harvard–Oxford subcortical ROIs, and (d) FreeSurfer brainstem subdivisions, all derived from the Severinghaus-derived SaO_2_ GLM. ICC values for the finer-grained 58-nucleus Brainstem Navigator atlas are shown in Supplemental Figure S3. For direct visual comparison, the corresponding 4-atlas ICC panels for the FIR, ET-trace, and SpO_2_ GLMs are provided in Supplemental Figures S8, S9, and S10, respectively; the four models yield qualitatively similar ICC patterns across atlases and hypercapnia/hypoxia conditions, with the SaO_2_ GLM showing modestly higher ROI-wise reproducibility overall. Within each panel, ROIs are ranked by ICC value; ROI label abbreviations are tabulated in the Supplemental Material.

**Figure 6:**
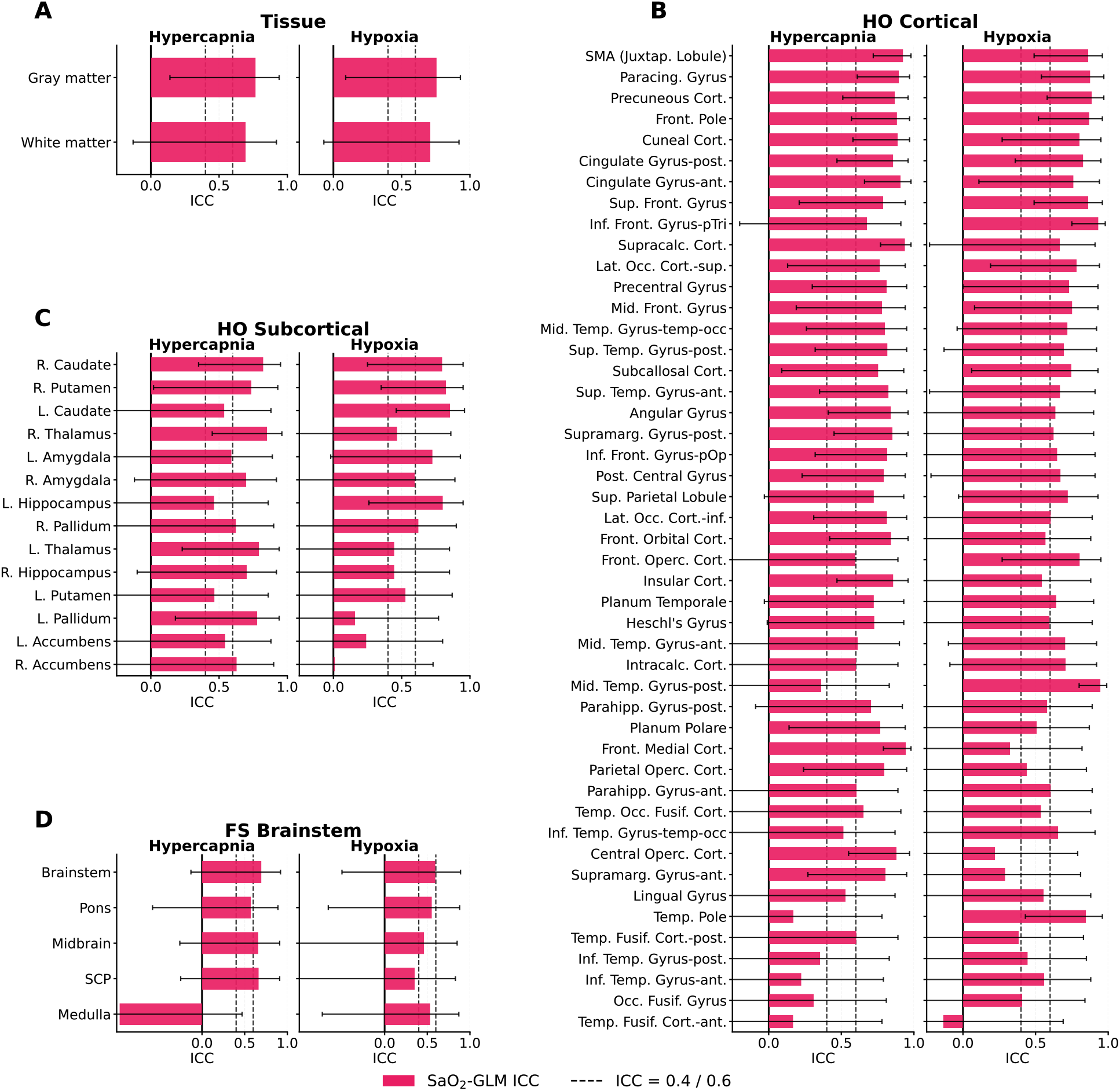
ROI-wise ICC of the Severinghaus-derived SaO_2_ GLM across four atlases at complementary spatial scales: (a) tissue compartments (gray matter, white matter), (b) Harvard–Oxford cortical ROIs, (c) Harvard–Oxford subcortical ROIs, and (d) FreeSurfer brainstem subdivisions. Within each panel, ROIs are ranked by mean ICC across hypercapnia and hypoxia (left and right sub-columns of each panel); vertical reference lines mark the ICC = 0.4 (fair) and 0.6 (good) reproducibility thresholds. ROI label abbreviations are tabulated in the Supplemental Material. Reproducibility is highest in tissue, cortical and subcortical ROIs (panels a, b, c) and drops progressively at finer brainstem scales (panel d), consistent with reduced BOLD SNR in the smaller, deeper structures where B0/B1 inhomogeneity and physiological noise are more pronounced.

## Discussion and Conclusions

### 4.1 Joint hypercapnic/hypoxic paradigm feasibility

A single 18-minute multi-echo, multi-band BOLD acquisition with a fixed-inspired continuous-flow gas manipulation yielded robust voxelwise hypercapnic and hypoxic CVR maps at 7 T. The fixed-inspired delivery produced ΔETCO_2_, ΔETO_2_, and ΔSpO_2_ amplitudes that were reproducible across sessions (ICCs 0.57–0.71; Table 1). Although fixed-inspired delivery affords less breath-by-breath control over the achieved end-tidal trajectory than prospective end-tidal targeting, the resulting gas changes were still large enough to elicit clearly modulated, condition-specific BOLD responses in both gray and white matter (Figure 2), and to support reproducible voxelwise CVR mapping under all four GLMs (Figure 5; FIR / ET-trace / SpO_2_ counterparts in Supplemental Figures S4–S6). Fixed-inspired delivery therefore remains a pragmatic CVR stimulus despite the loss of fine end-tidal control, consistent with three decades of prior use (Bandettini & Wong, 1997; Kastrup et al., 1999; Rostrup et al., 1995; Tancredi & Hoge, 2013).

### 4.2 Severinghaus-derived SaO_2_ regressor and hypoxia CVR

§1.6 identified the choice of hypoxia regressor as an unsettled question: the measured PETO_2_ trace, a saturation trace derived from PETO_2_ via Severinghaus- or Hill-style transformations, and the directly measured pulse-oximetry SpO_2_ trace each express hypoxic CVR in different physical units and rest on different assumptions, and to our knowledge no published study has compared the three within a single dataset. Our cross-session model-fit analysis (Supplemental Figure S2 and Figure 4) provides the first such comparison, and the result favours the Severinghaus-derived SaO_2_ trace: it generalizes significantly better than both the canonical end-tidal-trace GLM (sign-flip permutation *p* = 0.001) and the directly measured SpO_2_ GLM (*p* = 0.034) under two-fold cross-session validation. The trace-based SaO_2_ GLM was also less susceptible than the higher-degree-of-freedom FIR GLM to session-specific noise across 58% of gray-matter voxels — an expected consequence of the FIR’s greater flexibility, and one that illustrates the trade-off between within-session fit and cross-session generalization: the FIR’s per-time-point degrees of freedom fit the noise in the session on which it was estimated, and that within-session advantage does not translate directly into cross-session generalization.

Several factors likely explain the Severinghaus advantage. Physically, the BOLD signal during hypoxia is likely dominated by intravascular deoxyhemoglobin concentration (§1.4, §1.5) (Bhogal et al., 2022; Harris et al., 2013; Sayin et al., 2023), and [dOHb] is approximately proportional to 1 − SaO_2_ rather than to PETO_2_ in mmHg — so a Severinghaus-derived SaO_2_ trace is a closer physical proxy for the actual BOLD driver than the raw end-tidal trace it was derived from. The Severinghaus transform additionally incorporates a three-term Bohr correction that adjusts the derived SaO_2_ for instantaneous temperature, pH, and PETCO_2_ (Eq. 6); this matters specifically in our setting because, under fixed-inspired delivery, PETCO_2_ is **not** held constant — it is permitted to vary freely with each participant’s ventilatory response to the inhaled O_2_ and CO_2_ steps. A model that ignores the Bohr shift is misspecified in exactly the regime where ventilatory compensation is the largest source of inter-subject variability (a limitation that prospective end-tidal targeting (§1.3) circumvents by clamping PETCO_2_ to a prescribed trajectory). Statistically, the non-linear Severinghaus + Bohr transform amplifies subject-specific baseline-to-nadir saturation differences while suppressing breath-by-breath envelope noise in the raw ETO_2_ trace, consistent with the markedly higher across-session reproducibility of the derived ΔSaO_2_ amplitude (ICC = 0.95, 95% CI [0.87, 0.98]) than the underlying ΔETO_2_ amplitude (ICC = 0.57). Notably, although the recent perfusion-imaging lineage (Duffin et al., 2024; Sayin et al., 2023; Sayin et al., 2025; Stumpo et al., 2024) routinely uses the hypoxic step to drive DSC-style perfusion deconvolution, the saturation trace — where derived — is computed from PETO_2_ via the Balaban 2013 pH-dependent Hill-equation fit (Balaban et al., 2013) evaluated at a fixed baseline pH = 7.4, with no PCO_2_ or temperature correction; and it is routed into the arterial input function for perfusion deconvolution rather than into a BOLD-CVR GLM. Two methodological choices therefore distinguish the present work from that lineage: applying the full Severinghaus 1979 oxygen dissociation equation with the three-term Bohr correction for PCO_2_, pH, and temperature rather than the pH-only Balaban Hill fit, and using the resulting SaO_2_ trace as the primary BOLD GLM regressor rather than as a perfusion AIF — together providing a hypoxic counterpart to the canonical PETCO_2_-based hypercapnic CVR model.

### 4.3 Complementary role of the FIR GLM as a trace-free ΔBOLD estimator

Framed the other way, the FIR GLM’s independence from the recorded gas traces makes it a complementary tool rather than a competitor to the trace-based CVR models. Because the FIR fit does not use the end-tidal or peripheral-saturation traces as regressors, it is robust to measurement issues that can degrade trace-based fits — variable end-tidal trace quality (mask leaks, capnograph sampling delays), inter-subject differences in the achieved end-tidal trajectory under fixed-inspired delivery, and errors in temporally aligning each trace to the BOLD signal. The trade-off is that the raw FIR output is a per-condition %Δ BOLD amplitude rather than a CVR value normalized to the delivered end-tidal gas change: comparing FIR-derived %ΔBOLD magnitudes across subjects or across groups therefore risks confounding true differences in vascular reactivity with differences in the achieved gas dose, and normalization to ΔETCO_2_ or ΔETO_2_ remains necessary for such between-subject or between-group inference. Within a single subject, however, where the achieved gas dose is common across regions, the FIR-derived %ΔBOLD supports direct regional comparisons without gas-dose normalization — for example, contrasting the hypercapnic BOLD response magnitude across cortical, subcortical, and brainstem regions in the same individual. Supplemental Figure S7 shows that the FIR-derived %ΔBOLD achieves ROI-wise reproducibility comparable to the fully normalized CVR estimates: median ICC 0.79 (with 94% of ROI × condition tests reaching fair reproducibility, 80% good, and 59% excellent), essentially indistinguishable from the FIR-CVR variant (0.80; 97%, 90%, 67% respectively) despite skipping the gas-dose normalization step. The advantage of normalizing to the delivered gas dose is thus not one of reproducibility per se — it is one of physical interpretability and between-subject/between-group comparability. This supports the use of FIR-derived %ΔBOLD as a normalization-free CVR-like measure for within-subject applications where the gas dose is common across regions.

### 4.4 Multi-echo BOLD at 7T

The methodological choices reported here rely centrally on the multi-echo BOLD acquisition. First, optimal echo combination (DuPre et al., 2021) yields substantially improved signal-to-noise ratio and, more importantly at 7 T, recovers usable signal in high-susceptibility regions — orbitofrontal cortex, temporal poles, ventral cerebellum, and brainstem — where a single-echo acquisition at the same nominal resolution would lose signal to 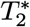 dropout. This is not incidental to our findings: the brain-wide reproducibility we report across cortical, subcortical, and brainstem atlases (§3.5, Figure 6) would not be achievable from a single-echo scan without either reducing spatial resolution or accepting large brainstem and orbitofrontal dropouts. Second, the voxelwise 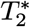 map produced during optimal combination provides an objective, signal-quality-based criterion for masking (§2.6): voxels with 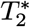 outside 10 to 40 ms — indicating either dropout or CSF/partial-volume contamination — are excluded from downstream ROI-mean CVR estimates without the need to hand-tune anatomical mask boundaries per subject. Third, the three-echo acquisition supports voxelwise 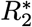 estimation, which we used to visualize the vascular response independently of the *S*_0_-modulated BOLD signal (Figure 2, bottom row) as an internal cross-check on the vascular origin of the observed BOLD excursions. In our view these three utilities — SNR/coverage, mask refinement, and 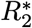 estimation — collectively make multi-echo BOLD the more prudent default than single-echo for gas-challenge CVR studies at 7 T, particularly when the study aims to characterize brainstem, deep gray-matter, or white-matter reactivity.

### 4.5 Reproducibility of hypercapnic and hypoxic CVR at 7 T

The second gap identified in §1.6 was the absence of any published test–retest characterization of gas-challenge CVR at 7 T, particularly for hypoxia. Our ROI-wise ICC analysis using the Severinghaus-derived SaO_2_ GLM (Figure 6; Brainstem Navigator ICCs in Supplemental Figure S3) provides the first such characterization: 74% of (ROI × condition) tests yielded fair-or-better reproducibility (ICC ≥ 0.4), 30% yielded excellent reproducibility (ICC ≥ 0.75), and the Harvard–Oxford cortical and subcortical ROIs reached a median ICC of 0.71. The lower reproducibility observed in the smaller Brainstem Navigator nuclei (median ICC = 0.49) is consistent with the well-known dependence of ICC on within-ROI BOLD SNR — small ROIs accumulate fewer voxels and inherit more thermal and physiological noise per measurement — and likely reflects a measurement-scale limitation rather than a true biological non-reproducibility of brainstem CVR. The brainstem is additionally a region in which BOLD SNR at 7 T is degraded by *B*_0_ and *B*_1_ inhomogeneity associated with its position deep in the head, near air–tissue interfaces, and far from the receive coil elements (Zuo et al., 2013), further depressing per-voxel reproducibility for the small nuclei resolved by the Brainstem Navigator atlas. Ongoing work in our group aims to optimize subject-specific *B*_0_ and *B*_1_ shimming protocols for brainstem-targeted 7 T BOLD, which we expect will recover a substantial portion of the reproducibility lost in this region. Together with the consistent voxelwise CVR map patterns across sessions (Figure 5), these results establish the combined hypercapnia + hypoxia paradigm as a reproducible probe of both axes of cerebrovascular reactivity at 7 T.

### 4.6 Limitations

The present study is limited by a small healthy-volunteer sample (*n* = 11) and a single 7 T site. We did not include a side-by-side comparison against a prospectively-targeted end-tidal forcing condition, leaving the absolute reproducibility cost of fixed-inspired vs end-tidal-forcing delivery as a question for future work. The Severinghaus 1979 oxygen dissociation equation with Bohr correction assumes population-typical baseline pH and temperature; subject-specific deviations from these assumptions may bias the derived SaO_2_ trace in clinical cohorts. We also did not acquire an independent ASL or spin-echo measurement that could directly partition the hypoxic BOLD response into deoxyhemoglobin and CBF/CBV contributions across the dose-and-duration regimes outlined in §1.4, leaving the relative contribution of flow vs deoxyhemoglobin to the observed hypoxic CVR maps unresolved within this dataset. Future work investigating dose-and-duration regimes on hypoxic BOLD response would contribute valuable insight by measuring CBF response dynamics and the reproducibility of such dynamics as well. Related to this, the 3-minute CBF-onset delay from Harris and colleagues (Harris et al., 2013) that motivates our short-bolus framing was measured under dynamic end-tidal forcing (which clamps PETO_2_ at a target trajectory breath-by-breath), whereas our paradigm delivers a fixed inspired O_2_ mixture and lets PETO_2_ follow each participant’s ventilatory response. The CBF response timing under fixed-inspired delivery has not been directly characterized and may differ modestly from Harris’s DEF-clamped estimate, but any such difference is unlikely to change the qualitative distinction between the transient-bolus and sustained-plateau regimes.

### 4.7 Conclusions

A continuous-flow, fixed-inspired hypercapnic + hypoxic gas paradigm at 7 T produces robust, reproducible BOLD CVR maps in a single 18-minute acquisition. Among the three physiologically motivated trace-based GLMs tested, a Severinghaus-derived SaO_2_ regressor – applied as a BOLD-CVR input – generalizes best across sessions and at the voxelwise level outperforms even the unconstrained FIR model, supporting its use as the default regressor for hypoxic CVR mapping. Together, the combined paradigm and the SaO_2_-GLM framework provide a feasible 7 T platform for probing two complementary axes of cerebrovascular reactivity, opening a path to applications in vascular ageing, cerebral small-vessel disease, and cerebrovascular reserve characterization in steno-occlusive disease.

## Data Availability

All data produced in the present study are available upon reasonable request to the authors.

## Declarations

### Ethics approval and consent to participate

All procedures were approved by the Faculty of Medicine and Health Sciences Research Ethics Board of McGill University (study number 23-07-074), and all relevant ethical guidelines were followed. All participants gave written informed consent prior to participation.

### Competing interests

The authors have declared no competing interests.

### Funding

This work was supported by the Canadian Institutes of Health Research (CIHR).

### Data availability

The datasets generated and analysed during the current study are available from the corresponding author on reasonable request.

## Supplemental Material

Supplemental tables and figures (Figures S1–S10, Tables S1–S4) are provided in a separate supplemental document.

## Notes

### Author Declarations

Ethics approval was obtained by the McGill Faculty of Medicine and Health Sciences REB, study 23-07-074.

## Bibliography

Abraham, A., Pedregosa, F., Eickenberg, M., Gervais, P., Mueller, A., Kossaifi, J., Gramfort, A., Thirion, B., & Varoquaux, G. (2014). Machine learning for neuroimaging with scikit-learn. Frontiers in Neuroinformatics, 8, 14. 10.3389/fninf.2014.00014

Andersson, J. L., Skare, S., & Ashburner, J. (2003). How to correct susceptibility distortions in spin-echo echo-planar images: application to diffusion tensor imaging. Neuroimage, 20(2), 870–888. 10.1016/S1053-8119(03)00336-7

Avants, B., Epstein, C., Grossman, M., & Gee, J. (2008). Symmetric diffeomorphic image registration with cross-correlation: Evaluating automated labeling of elderly and neurodegenerative brain. Medical Image Analysis, 12(1), 26–41. 10.1016/j.media.2007.06.004

Balaban, D. Y., Duffin, J., Preiss, D., Mardimae, A., Vesely, A., Slessarev, M., Zubieta-Calleja, G. R., Greene, E. R., MacLeod, D. B., & Fisher, J. A. (2013). The in-vivo oxyhaemoglobin dissociation curve at sea level and high altitude. Respiratory Physiology & Neurobiology, 186(1), 45–52. 10.1016/j.resp.2012.12.011

Bandettini, P. A., & Wong, E. C. (1997). A Hypercapnia-Based Normalization Method for Improved Spatial Localization of Human Brain Activation with fMRI. NMR in Biomedicine, 10(4–5), 197–203. 10.1002/(SICI)1099-1492(199706/08)10:4/5<197::AID-NBM466>3.0.CO;2-S

Bhogal, A. A., Philippens, M. E., Siero, J. C., Fisher, J. A., Petersen, E. T., Luijten, P. R., & Hoogduin, H. (2015). Examining the Regional and Cerebral Depth-Dependent BOLD Cerebrovascular Reactivity Response at 7 T. Neuroimage, 114, 239–248. 10.1016/j.neuroimage.2015.04.014

Bhogal, A. A., Sayin, E. S., Poublanc, J., Duffin, J., Fisher, J. A., Sobczyk, O., & Mikulis, D. J. (2022). Quantifying Cerebral Blood Arrival Times Using Hypoxia-Mediated Arterial BOLD Contrast. Neuroimage, 261, 119523. 10.1016/j.neuroimage.2022.119523

Blair, G. W., Doubal, F. N., Thrippleton, M. J., Marshall, I., & Wardlaw, J. M. (2016). Magnetic Resonance Imaging for Assessment of Cerebrovascular Reactivity in Cerebral Small Vessel Disease: A Systematic Review. Journal of Cerebral Blood Flow & Metabolism, 36(5), 833–841. 10.1177/0271678X16631756

Bright, M. G., & Murphy, K. (2013). Reliable Quantification of BOLD fMRI Cerebrovascular Reactivity despite Poor Breath-Hold Performance. Neuroimage, 83, 559–568. 10.1016/j.neuroimage.2013.07.007

Ciric, R., Thompson, W. H., Lorenz, R., Goncalves, M., MacNicol, E., Markiewicz, C. J., Halchenko, Y. O., Ghosh, S. S., Gorgolewski, K. J., Poldrack, R. A., & Esteban, O. (2022). TemplateFlow: FAIR-sharing of multi-scale, multi-species brain models. Nature Methods, 19, 1568–1571. 10.1038/s41592-022-01681-2

Dale, A. M., Fischl, B., & Sereno, M. I. (1999). Cortical Surface-Based Analysis: I. Segmentation and Surface Reconstruction. Neuroimage, 9(2), 179–194. 10.1006/nimg.1998.0395

Desikan, R. S., Ségonne, F., Fischl, B., Quinn, B. T., Dickerson, B. C., Blacker, D., Buckner, R. L., Dale, A. M., Maguire, R. P., Hyman, B. T., Albert, M. S., & Killiany, R. J. (2006). An Automated Labeling System for Subdividing the Human Cerebral Cortex on MRI Scans into Gyral Based Regions of Interest. Neuroimage, 31(3), 968–980. 10.1016/j.neuroimage.2006.01.021

Duffin, J., Sayin, E. S., Sobczyk, O., Poublanc, J., Mikulis, D. J., & Fisher, J. A. (2024). Cerebral perfusion metrics calculated directly from a hypoxia-induced step change in deoxyhemoglobin. Scientific Reports, 14(1), 17121. 10.1038/s41598-024-68047-w

DuPre, E., Salo, T., Ahmed, Z., Bandettini, P. A., Bottenhorn, K. L., Caballero-Gaudes, C., Dowdle, L. T., Gonzalez-Castillo, J., Heunis, S., Kundu, P., Laird, A. R., Markello, R., Markiewicz, C. J., Moia, S., Staden, I., Teves, J. B., Uruñuela, E., Vaziri-Pashkam, M., Whitaker, K., & Handwerker, D. A. (2021). TE-dependent analysis of multi-echo fMRI with tedana. Journal of Open Source Software, 6(66), 3669. 10.21105/joss.03669

Esteban, O., Blair, R., Markiewicz, C. J., Berleant, S. L., Moodie, C., Ma, F., Isik, A. I., Erramuzpe, A., Kent, M., James D. and Goncalves DuPre, E., Sitek, K. R., Gomez, D. E. P., Lurie, D. J., Ye, Z., Poldrack, R. A., & Gorgolewski, K. J. (2018). fMRIPrep <version>. Software. 10.5281/zenodo.852659

Esteban, O., Markiewicz, C., Blair, R. W., Moodie, C., Isik, A. I., Erramuzpe Aliaga, A., Kent, J., Goncalves, M., DuPre, E., Snyder, M., Oya, H., Ghosh, S., Wright, J., Durnez, J., Poldrack, R., & Gorgolewski, K. J. (2019). fMRIPrep: a robust preprocessing pipeline for functional MRI. Nature Methods, 16, 111–116. 10.1038/s41592-018-0235-4

Fonov, V., Evans, A., McKinstry, R., Almli, C., & Collins, D. (2009). Unbiased nonlinear average age-appropriate brain templates from birth to adulthood. Neuroimage, 47, Supplement 1, S102. 10.1016/S1053-8119(09)70884-5

Glover, G. H. (1999). Deconvolution of impulse response in event-related BOLD fMRI. Neuroimage, 9(4), 416–429. 10.1006/nimg.1998.0419

Gorgolewski, K., Burns, C. D., Madison, C., Clark, D., Halchenko, Y. O., Waskom, M. L., & Ghosh, S. (2011). Nipype: a flexible, lightweight and extensible neuroimaging data processing framework in Python. Frontiers in Neuroinformatics, 5, 13. 10.3389/fninf.2011.00013

Gorgolewski, K. J., Esteban, O., Markiewicz, C. J., Ziegler, E., Ellis, D. G., Notter, M. P., Jarecka, D., Johnson, H., Burns, C., Manhães-Savio, A., Hamalainen, C., Yvernault, B., Salo, T., Jordan, K., Goncalves, M., Waskom, M., Clark, D., Wong, J., Loney, F., … Ghosh, S. (2018). Nipype. Software. 10.5281/zenodo.596855

Greve, D. N., & Fischl, B. (2009). Accurate and robust brain image alignment using boundary-based registration. Neuroimage, 48(1), 63–72. 10.1016/j.neuroimage.2009.06.060

Hansen, J. Y., Cauzzo, S., Singh, K., García-Gomar, M. G., Shine, J. M., Bianciardi, M., & Misic, B. (2024). Integrating Brainstem and Cortical Functional Architectures. Nature Neuroscience, 27(12), 2500–2511. 10.1038/s41593-024-01787-0

Harris, A. D., Murphy, K., Diaz, C. M., Saxena, N., Hall, J. E., Liu, T. T., & Wise, R. G. (2013). Cerebral Blood Flow Response to Acute Hypoxic Hypoxia. NMR in Biomedicine, 26(12), 1844–1852. 10.1002/nbm.3026

Hoge, R. D., Atkinson, J., Gill, B., Crelier, G. R., Marrett, S., & Pike, G. B. (1999). Investigation of BOLD signal dependence on cerebral blood flow and oxygen consumption: the deoxyhemoglobin dilution model. Magnetic Resonance in Medicine, 42(5), 849–863. 10.1002/(SICI)1522-2594(199911)42:5<849::AID-MRM4>3.0.CO;2-Z

Jefferson, A. L., Cambronero, F. E., Liu, D., Moore, E. E., Neal, J. E., Terry, J. G., Nair, S., Pechman, K. R., Rane, S., Davis, L. T., Gifford, K. A., Hohman, T. J., Bell, S. P., Wang, T. J., Beckman, J. A., & Carr, J. J. (2018). Higher Aortic Stiffness Is Related to Lower Cerebral Blood Flow and Preserved Cerebrovascular Reactivity in Older Adults. Circulation, 138(18), 1951–1962. 10.1161/CIRCULATIONAHA.118.032410

Jenkinson, M., & Smith, S. (2001). A global optimisation method for robust affine registration of brain images. Medical Image Analysis, 5(2), 143–156. 10.1016/S1361-8415(01)00036-6

Jenkinson, M., Beckmann, C. F., Behrens, T. E., Woolrich, M. W., & Smith, S. M. (2012). FSL. Neuroimage, 62(2), 782–790. 10.1016/j.neuroimage.2011.09.015

Jezzard, P., Heineman, F., Taylor, J., DesPres, D., Wen, H., Balaban, R. S., & Turner, R. (1994). Comparison of EPI Gradient-Echo Contrast Changes in Cat Brain Caused by Respiratory Challenges with Direct Simultaneous Evaluation of Cerebral Oxygenation via a Cranial Window. NMR in Biomedicine, 7(1– 2), 35–44. 10.1002/nbm.1940070107

Kastrup, A., Krüger, G., Glover, G. H., Neumann-Haefelin, T., & Moseley, M. E. (1999). Regional Variability of Cerebral Blood Oxygenation Response to Hypercapnia. Neuroimage, 10(6), 675–681. 10.1006/nimg.1999.0505

Keeling, E. G., Bergamino, M., Ott, L. R., McElvogue, M. M., & Stokes, A. M. (2025). Repeatability and Reliability of Cerebrovascular Reactivity in Young Adults Using Multi-Echo, Multi-Contrast MRI. Journal of Cerebral Blood Flow & Metabolism, 45(10), 2030–2046. 10.1177/0271678X251345292

Klein, A., Ghosh, S. S., Bao, F. S., Giard, J., Häme, Y., Stavsky, E., Lee, N., Rossa, B., Reuter, M., Neto, E. C., & Keshavan, A. (2017). Mindboggling morphometry of human brains. PLOS Computational Biology, 13(2), e1005350. 10.1371/journal.pcbi.1005350

Koo, T. K., & Li, M. Y. (2016). A Guideline of Selecting and Reporting Intraclass Correlation Coefficients for Reliability Research. Journal of Chiropractic Medicine, 15(2), 155–163. 10.1016/j.jcm.2016.02.012

Krishnamurthy, V., Sprick, J. D., Krishnamurthy, L. C., Barter, J. D., Turabi, A., Hajjar, I. M., & Nocera, J. R. (2021). The Utility of Cerebrovascular Reactivity MRI in Brain Rehabilitation: A Mechanistic Perspective. Frontiers in Physiology, 12, 642850. 10.3389/fphys.2021.642850

Liu, P., De Vis, J. B., & Lu, H. (2019). Cerebrovascular Reactivity (CVR) MRI with CO2 Challenge: A Technical Review. Neuroimage, 187, 104–115. 10.1016/j.neuroimage.2018.03.047

Liu, P., Ernst, T., Liang, H., Jiang, D., Cunningham, E., Ryan, M., Lu, H., Kottilil, S., & Chang, L. (2024). Elevated cerebral oxygen extraction in patients with post-COVID conditions. Neuroimmune Pharmacology and Therapeutics, 3(3–4), 169–174. 10.1515/nipt-2024-0014

Lu, H., Xu, F., Rodrigue, K. M., Kennedy, K. M., Cheng, Y., Flicker, B., Hebrank, A. C., Uh, J., & Park, D. C. (2011). Alterations in Cerebral Metabolic Rate and Blood Supply across the Adult Lifespan. Cerebral Cortex, 21(6), 1426–1434. 10.1093/cercor/bhq224

Makris, N., Goldstein, J. M., Kennedy, D., Hodge, S. M., Caviness, V. S., Faraone, S. V., Tsuang, M. T., & Seidman, L. J. (2006). Decreased Volume of Left and Total Anterior Insular Lobule in Schizophrenia. Schizophrenia Research, 83(2–3), 155–171. 10.1016/j.schres.2005.11.020

Mandell, D. M., Han, J. S., Poublanc, J., Crawley, A. P., Stainsby, J. A., Fisher, J. A., & Mikulis, D. J. (2008). Mapping Cerebrovascular Reactivity Using Blood Oxygen Level-Dependent MRI in Patients With Arterial Steno-occlusive Disease: Comparison With Arterial Spin Labeling MRI. Stroke, 39(7), 2021–2028. 10.1161/STROKEAHA.107.506709

McKetton, L., Venkatraghavan, L., Rosen, C., Mandell, D., Sam, K., Sobczyk, O., Poublanc, J., Gray, E., Crawley, A., Duffin, J., Fisher, J., & Mikulis, D. (2019). Improved White Matter Cerebrovascular Reactivity after Revascularization in Patients with Steno-Occlusive Disease. American Journal of Neuroradiology, 40(1), 45–50. 10.3174/ajnr.A5912

Mitsis, G. D., Poulin, M. J., Robbins, P. A., & Marmarelis, V. Z. (2004). Nonlinear modeling of the dynamic effects of arterial pressure and CO2 variations on cerebral blood flow in healthy humans. IEEE Transactions on Biomedical Engineering, 51(11), 1932–1943. 10.1109/TBME.2004.834272

Ogawa, Lee Kay, et al. (1990). Brain Magnetic Resonance Imaging with Contrast Dependent on Blood Oxygenation. Proceedings of the National Academy of Sciences, 87(24), 9868–9872. 10.1073/pnas.87.24.9868

Ogawa, Lee Nayak, et al. (1990). Oxygenation-Sensitive Contrast in Magnetic Resonance Image of Rodent Brain at High Magnetic Fields. Magnetic Resonance in Medicine, 14(1), 68–78. 10.1002/mrm.1910140108

Prielmeier, F., Nagatomo, Y., & Frahm, J. (1994). Cerebral Blood Oxygenation in Rat Brain during Hypoxic Hypoxia. Quantitative MRI of Effective Transverse Relaxation Rates. Magnetic Resonance in Medicine, 31(6), 678–681. 10.1002/mrm.1910310615

Prokopiou, P. C., Pattinson, K. T., Wise, R. G., & Mitsis, G. D. (2019). Modeling of Dynamic Cerebrovascular Reactivity to Spontaneous and Externally Induced CO2 Fluctuations in the Human Brain Using BOLD-fMRI. Neuroimage, 186, 533–548. 10.1016/j.neuroimage.2018.10.084

Reuter, M., Rosas, H. D., & Fischl, B. (2010). Highly accurate inverse consistent registration: A robust approach. Neuroimage, 53(4), 1181–1196. 10.1016/j.neuroimage.2010.07.020

Rostrup, E., Larsson, H. B. W., Toft, P. B., Garde, K., & Henriksen, O. (1995). Signal Changes in Gradient Echo Images of Human Brain Induced by Hypo- and Hyperoxia. NMR in Biomedicine, 8(1), 41–47. 10.1002/nbm.1940080109

Sayin, E. S., Duffin, J., Poublanc, J., Fisher, J. A., Mikulis, D. J., & Sobczyk, O. (2025). MRI-Based Classification of Cerebral Hemodynamic Failure With Resting Perfusion Metrics and Cerebrovascular Reactivity. Stroke, 56(10), 3034–3046. 10.1161/STROKEAHA.125.050978

Sayin, E. S., Schulman, J., Poublanc, J., Levine, H. T., Venkat Raghavan, L., Uludag, K., Duffin, J., Fisher, J. A., Mikulis, D. J., & Sobczyk, O. (2023). Investigations of Hypoxia-Induced Deoxyhemoglobin as a Contrast Agent for Cerebral Perfusion Imaging. Human Brain Mapping, 44(3), 1019–1029. 10.1002/hbm.26131

Severinghaus, J. W. (1979). Simple, Accurate Equations for Human Blood O2 Dissociation Computations. Journal of Applied Physiology, 46(3), 599–602. 10.1152/jappl.1979.46.3.599

Shams, S., Prokopiou, P., Esmaelbeigi, A., Mitsis, G. D., & Chen, J. J. (2023). Modeling the dynamics of cerebrovascular reactivity to carbon dioxide in fMRI under task and resting-state conditions. Neuroimage, 265, 119758. 10.1016/j.neuroimage.2022.119758

Shrout, P. E., & Fleiss, J. L. (1979). Intraclass Correlations: Uses in Assessing Rater Reliability. Psychological Bulletin, 86(2), 420–428.

Slessarev, M., Han, J., Mardimae, A., Prisman, E., Preiss, D., Volgyesi, G., Ansel, C., Duffin, J., & Fisher, J. A. (2007). Prospective Targeting and Control of End-Tidal CO\textsubscript{2} and O\textsubscript{2} Concentrations. The Journal of Physiology, 581(3), 1207–1219. 10.1113/jphysiol.2007.129395

Stehling, M. K., Schmitt, F., & Ladebeck, R. (1993). Echo-Planar MR Imaging of Human Brain Oxygenation Changes. Journal of Magnetic Resonance Imaging, 3(2), 471–474. 10.1002/jmri.1880030307

Stumpo, V., Sayin, E. S., Bellomo, J., Sobczyk, O., Van Niftrik, C. H. B., Sebök, M., Weller, M., Regli, L., Kulcsár, Z., Pangalu, A., Bink, A., Duffin, J., Mikulis, D. D., Fisher, J. A., & Fierstra, J. (2024). Transient deoxyhemoglobin formation as a contrast for perfusion MRI studies in patients with brain tumors: a feasibility study. Frontiers in Physiology, 15, 1238533. 10.3389/fphys.2024.1238533

Switzer, A. R., Cheema, I., McCreary, C. R., Zwiers, A., Charlton, A., Alvarez-Veronesi, A., Sekhon, R., Zerna, C., Stafford, R. B., Frayne, R., Goodyear, B. G., & Smith, E. E. (2020). Cerebrovascular Reactivity in Cerebral Amyloid Angiopathy, Alzheimer Disease, and Mild Cognitive Impairment. Neurology, 95(10). 10.1212/WNL.0000000000010201

Tancredi, F. B., & Hoge, R. D. (2013). Comparison of Cerebral Vascular Reactivity Measures Obtained Using Breath-Holding and CO\textsubscript{2} Inhalation. Journal of Cerebral Blood Flow & Metabolism, 33(7), 1066–1074. 10.1038/jcbfm.2013.48

Thomas, B. P., Liu, P., Park, D. C., Van Osch, M. J., & Lu, H. (2014). Cerebrovascular Reactivity in the Brain White Matter: Magnitude, Temporal Characteristics, and Age Effects. Journal of Cerebral Blood Flow & Metabolism, 34(2), 242–247. 10.1038/jcbfm.2013.194

Thomas, B. P., Liu, P., Aslan, S., King, K. S., Van Osch, M. J., & Lu, H. (2013). Physiologic Underpinnings of Negative BOLD Cerebrovascular Reactivity in Brain Ventricles. Neuroimage, 83, 505–512. 10.1016/j.neuroimage.2013.07.005

Turner, R., Le Bihan, D., Moonen, C. T. W., Despres, D., & Frank, J. (1991). Echo-Planar Time Course MRI of Cat Brain Oxygenation Changes. Magnetic Resonance in Medicine, 22(1), 159–166. 10.1002/mrm.1910220117

Tustison, N. J., Avants, B. B., Cook, P. A., Zheng, Y., Egan, A., Yushkevich, P. A., & Gee, J. C. (2010). N4ITK: Improved N3 Bias Correction. IEEE Transactions on Medical Imaging, 29(6), 1310–1320. 10.1109/TMI.2010.2046908

Vallat, R. (2018). Pingouin: Statistics in Python. Journal of Open Source Software, 3(31), 1026. 10.21105/joss.01026

Vorstrup, S., Brun, B., & Lassen, N. A. (1986). Evaluation of the Cerebral Vasodilatory Capacity by the Acetazolamide Test before EC-IC Bypass Surgery in Patients with Occlusion of the Internal Carotid Artery. Stroke, 17(6), 1291–1298. 10.1161/01.STR.17.6.1291

Wise, R. G., Ide, K., Poulin, M. J., & Tracey, I. (2004). Resting fluctuations in arterial carbon dioxide induce significant low frequency variations in BOLD signal. Neuroimage, 21(4), 1652–1664. 10.1016/j.neuroimage.2003.11.025

Wise, R. G., Pattinson, K. T. S., Bulte, D. P., Chiarelli, P. A., Mayhew, S. D., Balanos, G. M., O’Connor, D. F., Pragnell, T. R., Robbins, P. A., Tracey, I., & Jezzard, P. (2007). Dynamic Forcing of End-Tidal Carbon Dioxide and Oxygen Applied to Functional Magnetic Resonance Imaging. Journal of Cerebral Blood Flow & Metabolism, 27(8), 1521–1532. 10.1038/sj.jcbfm.9600465

Wise, R. G., Pattinson, K. T. S., Bulte, D. P., Rogers, R., Tracey, I., Matthews, P. M., & Jezzard, P. (2010). Measurement of Relative Cerebral Blood Volume Using BOLD Contrast and Mild Hypoxic Hypoxia. Magnetic Resonance Imaging, 28(8), 1129–1134. 10.1016/j.mri.2010.06.002

Wittich, W., Phillips, N., Nasreddine, Z. S., & Chertkow, H. (2010). Sensitivity and specificity of the Montreal Cognitive Assessment modified for individuals who are visually impaired. Journal of Visual Impairment & Blindness, 104(6), 360–368. 10.1177/0145482X1010400606

Yezhuvath, U. S., Uh, J., Cheng, Y., Martin-Cook, K., Weiner, M., Diaz-Arrastia, R., Van Osch, M., & Lu, H. (2012). Forebrain-Dominant Deficit in Cerebrovascular Reactivity in Alzheimer’s Disease. Neurobiology of Aging, 33(1), 75–82. 10.1016/j.neurobiolaging.2010.02.005

Zhang, Y., Brady, M., & Smith, S. (2001). Segmentation of brain MR images through a hidden Markov random field model and the expectation-maximization algorithm. IEEE Transactions on Medical Imaging, 20(1), 45–57. 10.1109/42.906424

Zuo, Z., Wang, R., Zhuo, Y., Xue, R., Lawrence, K. S. S., & Wang, D. J. J. (2013). Turbo-FLASH Based Arterial Spin Labeled Perfusion MRI at 7 T. Plos ONE, 8(6), e66612. 10.1371/journal.pone.0066612

