## Supplemental_Materials for "Cerebrovascular Reactivity to Hypoxic and Hypercapnic Gas Challenges Using 7T BOLD fMRI"

### Supplemental Tables: ROI Abbreviations

The ranked ICC figures (main-text Figure 6, panels b–d, and Supplemental Figure S3) display ROI labels in abbreviated form to keep axis tick labels readable. The following tables provide the full label-to-abbreviation mapping for each atlas.

Table S1: Harvard–Oxford cortical ROI label → abbreviation mapping (used in main-text Figure 6, panel b).

| Index | Label | Abbreviation |
|---|---|---|
| 1 | Frontal-Pole | Front. Pole |
| 2 | Insular-Cortex | Insular Cort. |
| 3 | Superior-Frontal-Gyrus | Sup. Front. Gyrus |
| 4 | Middle-Frontal-Gyrus | Mid. Front. Gyrus |
| 5 | Inferior-Frontal-Gyrus--pars-triangularis | Inf. Front. Gyrus-pTri |
| 6 | Inferior-Frontal-Gyrus--pars-opercularis | Inf. Front. Gyrus-pOp |
| 7 | Precentral-Gyrus | Precentral Gyrus |
| 8 | Temporal-Pole | Temp. Pole |
| 9 | Superior-Temporal-Gyrus--anterior-division | Sup. Temp. Gyrus-ant. |
| 10 | Superior-Temporal-Gyrus--posterior-division | Sup. Temp. Gyrus-post. |
| 11 | Middle-Temporal-Gyrus--anterior-division | Mid. Temp. Gyrus-ant. |
| 12 | Middle-Temporal-Gyrus--posterior-division | Mid. Temp. Gyrus-post. |
| 13 | Middle-Temporal-Gyrus--temporooccipital-part | Mid. Temp. Gyrus-temp-occ |
| 14 | Inferior-Temporal-Gyrus--anterior-division | Inf. Temp. Gyrus-ant. |
| 15 | Inferior-Temporal-Gyrus--posterior-division | Inf. Temp. Gyrus-post. |
| 16 | Inferior-Temporal-Gyrus--temporooccipital-part | Inf. Temp. Gyrus-temp-occ |
| 17 | Postcentral-Gyrus | Post. Central Gyrus |
| 18 | Superior-Parietal-Lobule | Sup. Parietal Lobule |
| 19 | Supramarginal-Gyrus--anterior-division | Supramarg. Gyrus-ant. |
| 20 | Supramarginal-Gyrus--posterior-division | Supramarg. Gyrus-post. |
| 21 | Angular-Gyrus | Angular Gyrus |
| 22 | Lateral-Occipital-Cortex--superior-division | Lat. Occ. Cort.-sup. |
| 23 | Lateral-Occipital-Cortex--inferior-division | Lat. Occ. Cort.-inf. |
| 24 | Intracalcarine-Cortex | Intracalc. Cort. |
| 25 | Frontal-Medial-Cortex | Front. Medial Cort. |
| 26 | Juxtapositional-Lobule-Cortex-(formerly-Supplementary-Motor-Cortex) | SMA (Juxtap. Lobule) |
| 27 | Subcallosal-Cortex | Subcallosal Cort. |
| 28 | Paracingulate-Gyrus | Paracing. Gyrus |
| 29 | Cingulate-Gyrus--anterior-division | Cingulate Gyrus-ant. |
| 30 | Cingulate-Gyrus--posterior-division | Cingulate Gyrus-post. |
| 31 | Precuneous-Cortex | Precuneous Cort. |
| 32 | Cuneal-Cortex | Cuneal Cort. |
| 33 | Frontal-Orbital-Cortex | Front. Orbital Cort. |
| [illegible] | [illegible] | [illegible] |

Table S2: Harvard–Oxford subcortical ROI label → abbreviation mapping (used in main-text Figure 6, panel c).

| **Label** | **Abbreviation** |
|---|---|
| Left-Cerebral-White-Matter | L. Cerebral WM |
| Left-Cerebral-Cortex | L. Cerebral Cortex |
| Left-Lateral-Ventricle | L. Lat. Ventricle |
| Left-Thalamus | L. Thalamus |
| Left-Caudate | L. Caudate |
| Left-Putamen | L. Putamen |
| Left-Pallidum | L. Pallidum |
| Brain-Stem | Brainstem |
| Left-Hippocampus | L. Hippocampus |
| Left-Amygdala | L. Amygdala |
| Left-Accumbens | L. Accumbens |
| Right-Cerebral-White-Matter | R. Cerebral WM |
| Right-Cerebral-Cortex | R. Cerebral Cortex |
| Right-Lateral-Ventricle | R. Lat. Ventricle |
| Right-Thalamus | R. Thalamus |
| Right-Caudate | R. Caudate |
| Right-Putamen | R. Putamen |
| Right-Pallidum | R. Pallidum |
| Right-Hippocampus | R. Hippocampus |
| Right-Amygdala | R. Amygdala |
| Right-Accumbens | R. Accumbens |

Table S3: FreeSurfer brainstem subdivision label → abbreviation mapping (used in main-text Figure 6, panel d).

| **Label** | **Abbreviation** |
|---|---|
| brainstem_combined | Brainstem (combined) |
| brainstem_pons | Pons |
| brainstem_midbrain | Midbrain |
| brainstem_scp | Sup. Cerebell. Peduncle |
| brainstem_medulla | Medulla |

Table S4: Brainstem Navigator nucleus label → abbreviation mapping (used in main-text Figure S3).

| **Label** | **Abbreviation** |
| --- | --- |
| CLi_RLi | Caud./Rostr. Linear Raphe |
| CnF | Cuneiform |
| CnF_l | Cuneiform (L) |
| CnF_r | Cuneiform (R) |
| DR | Dorsal Raphe |
| IC_l | Inf. Colliculus (L) |
| IC_r | Inf. Colliculus (R) |
| ION_l | Inf. Olivary (L) |
| ION_r | Inf. Olivary (R) |
| LC_l | Loc. Coer. (L) |
| LC_r | Loc. Coer. (R) |
| LDTg_CGPn_l | Latdors. Tegm./CGPn (L) |
| LDTg_CGPn_r | Latdors. Tegm./CGPn (R) |
| LPB_l | Lat. Parabrach. (L) |
| LPB_r | Lat. Parabrach. (R) |
| MPB_l | Med. Parabrach. (L) |
| MPB_r | Med. Parabrach. (R) |
| MiTg_PBG_l | MiTg/Parabigem. (L) |
| MiTg_PBG_r | MiTg/Parabigem. (R) |
| MnR | Median Raphe |
| PAG | Periaq. Gray |
| PCRtA_l | Parvoc. Retic. α (L) |
| PCRtA_r | Parvoc. Retic. α (R) |
| PMnR | Paramed. Raphe |
| PTg_l | Peduncul. Tegm. (L) |
| PTg_r | Peduncul. Tegm. (R) |
| PnO_PnC_l | Pontine O/C (L) |
| PnO_PnC_r | Pontine O/C (R) |
| RMg | Raphe Magnus |
| RN_l | Red Nucleus (L) |
| RN_r | Red Nucleus (R) |
| RN1_l | Red Nucl. 1 (L) |
| RN1_r | Red Nucl. 1 (R) |
| [illegible] | [illegible] |

### Supplemental Figures: GLM Model-Fit Comparison

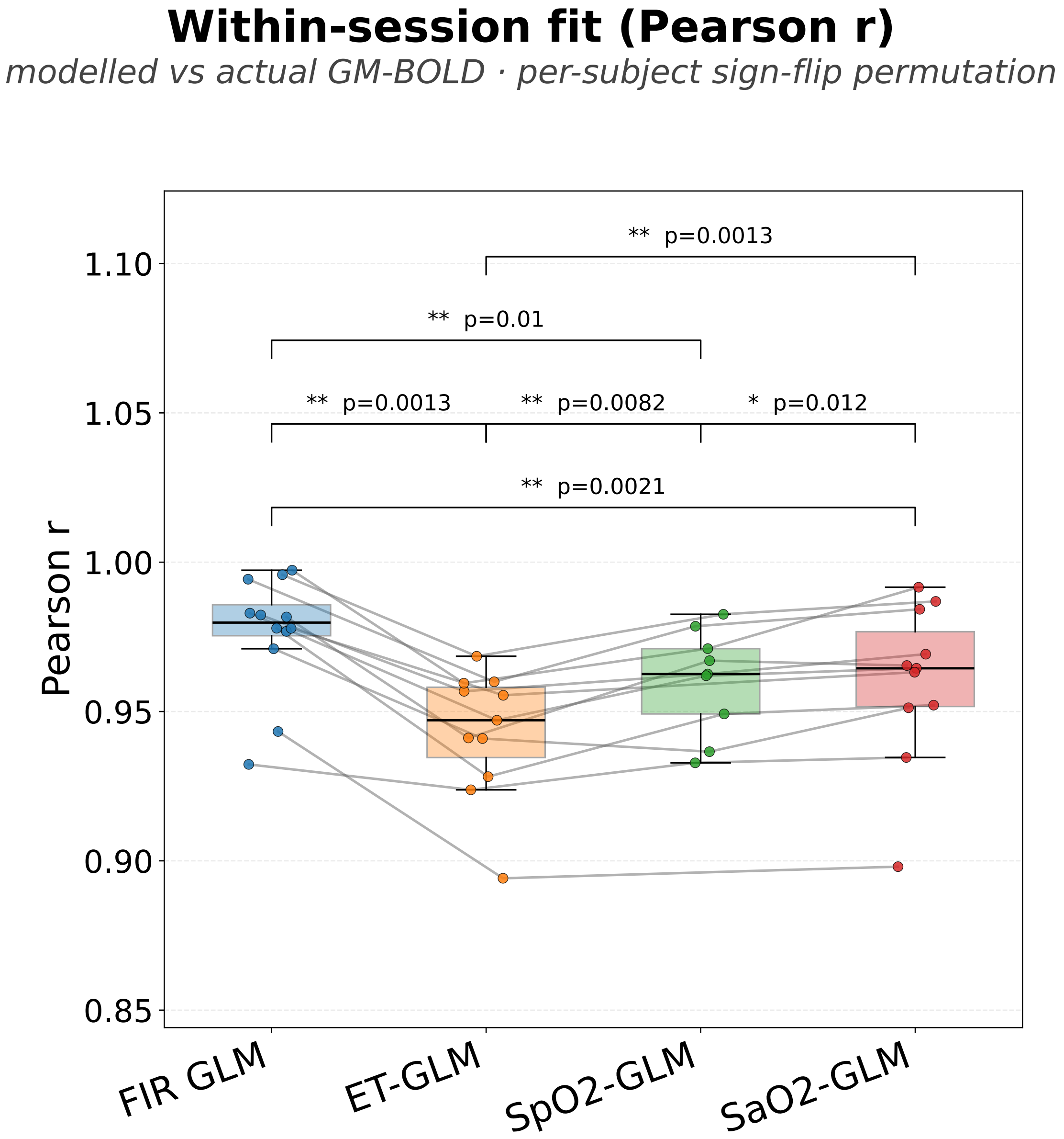


Figure S1: Within-session Pearson $r$ between predicted and observed gray-matter BOLD time-series, compared across the four GLMs (FIR, ET-trace, $SpO_2$, Severinghaus-derived $SaO_2$). Each point is a single participant; brackets denote pairwise contrasts with significant sign-flip permutation tests (10,000 permutations). The FIR GLM significantly outperformed every trace-based model within-session, reflecting its much larger degrees of freedom; this advantage does not persist under cross-session validation (Supplemental Figure S2).

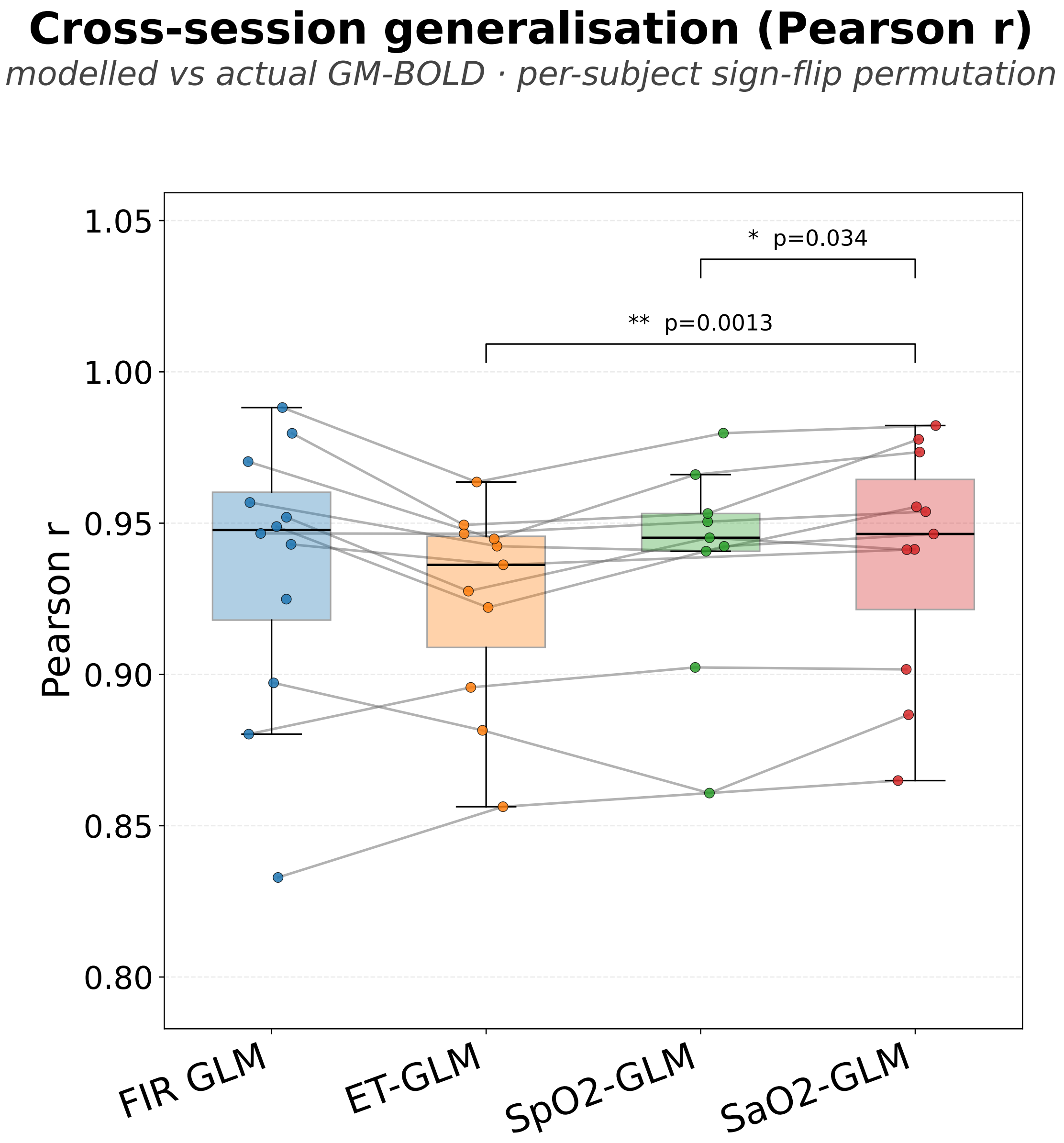


Figure S2: Cross-session Pearson $r$ between predicted and observed gray-matter BOLD time-series, compared across the four GLMs (FIR, ET-trace, $SpO_2$, Severinghaus-derived $SaO_2$). Each point is a single participant; brackets denote pairwise contrasts with significant sign-flip permutation tests (10,000 permutations). Under cross-session validation the $SaO_2$ GLM was the only model with pairwise advantages that survived permutation testing, generalizing significantly better than both the ET-GLM (permutation $p = 0.001$) and the $SpO_2$ GLM ($p = 0.034$); the FIR GLM's within-session advantage (Supplemental Figure S1) did not persist across sessions.

### Supplemental Figures: Brainstem Navigator ICC

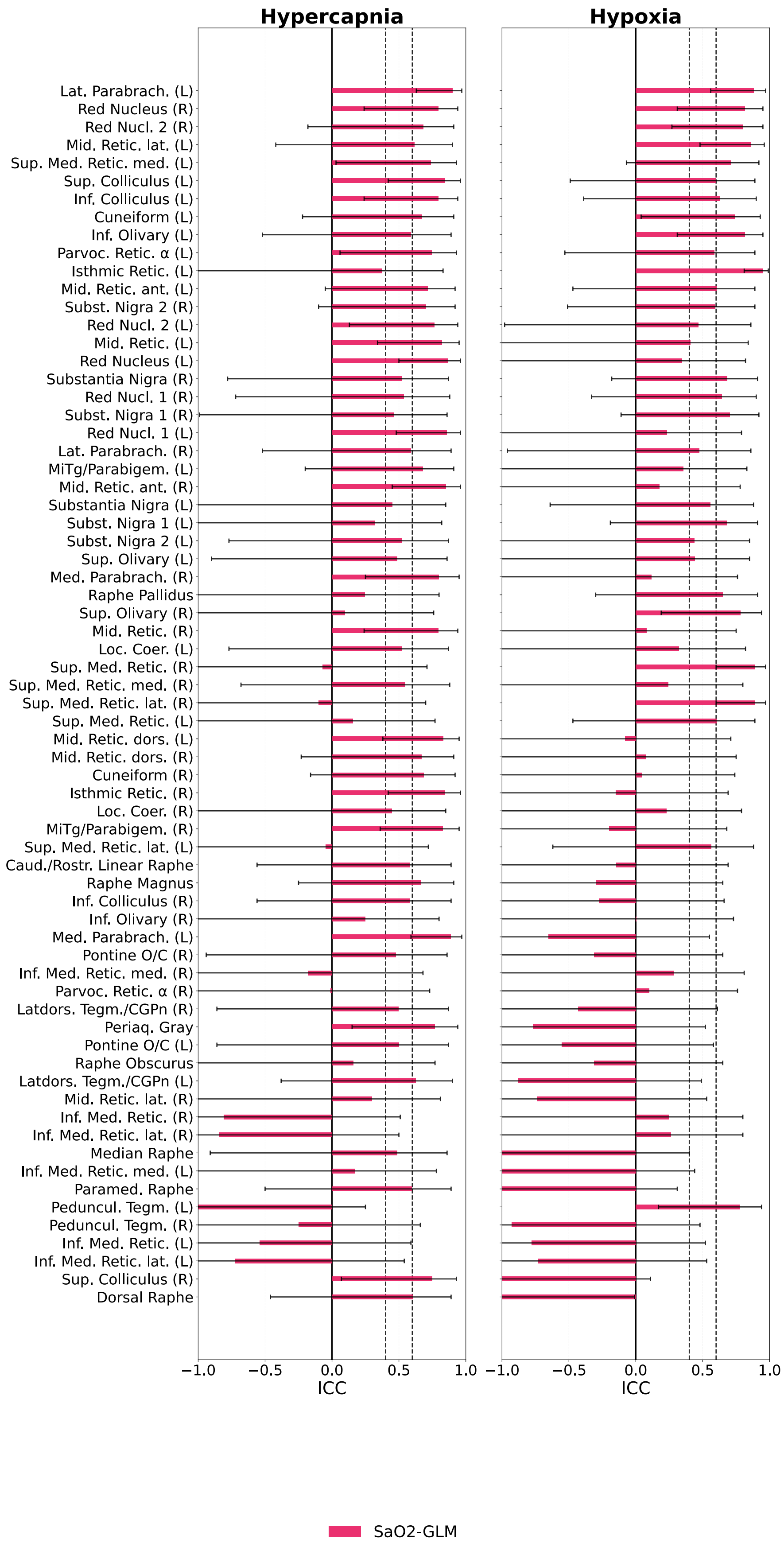


Figure S3: ICC of the Severinghaus-derived $SaO_2$ GLM for the 58-nucleus Brainstem Navigator atlas, ranked by ICC value. Median ICC across nuclei is 0.49, with 37% of (nucleus × condition) tests achieving ICC > 0.6 — lower than the coarser FreeSurfer brainstem subdivisions shown in main-text Figure 12, consistent with the smaller voxel counts per nucleus and the reduced BOLD SNR at 7 T in the brainstem [illegible]

### Supplemental Figures: Voxelwise CVR Maps for FIR, ET-trace, and $SpO_2$ GLMs

The main text presents voxelwise CVR maps derived from the Severinghaus-derived $SaO_2$ GLM (main-text Figure 8), which was identified as the most reproducible and best-generalizing model of the gas-induced BOLD response (main-text §3.3). The corresponding group-mean CVR maps derived from the FIR, ET-trace, and $SpO_2$ GLMs are provided below for direct comparison. All four GLMs produce qualitatively similar hypercapnic (top two rows) and hypoxic (bottom two rows) CVR patterns, with the same gray-matter spatial coverage and session-to-session consistency.

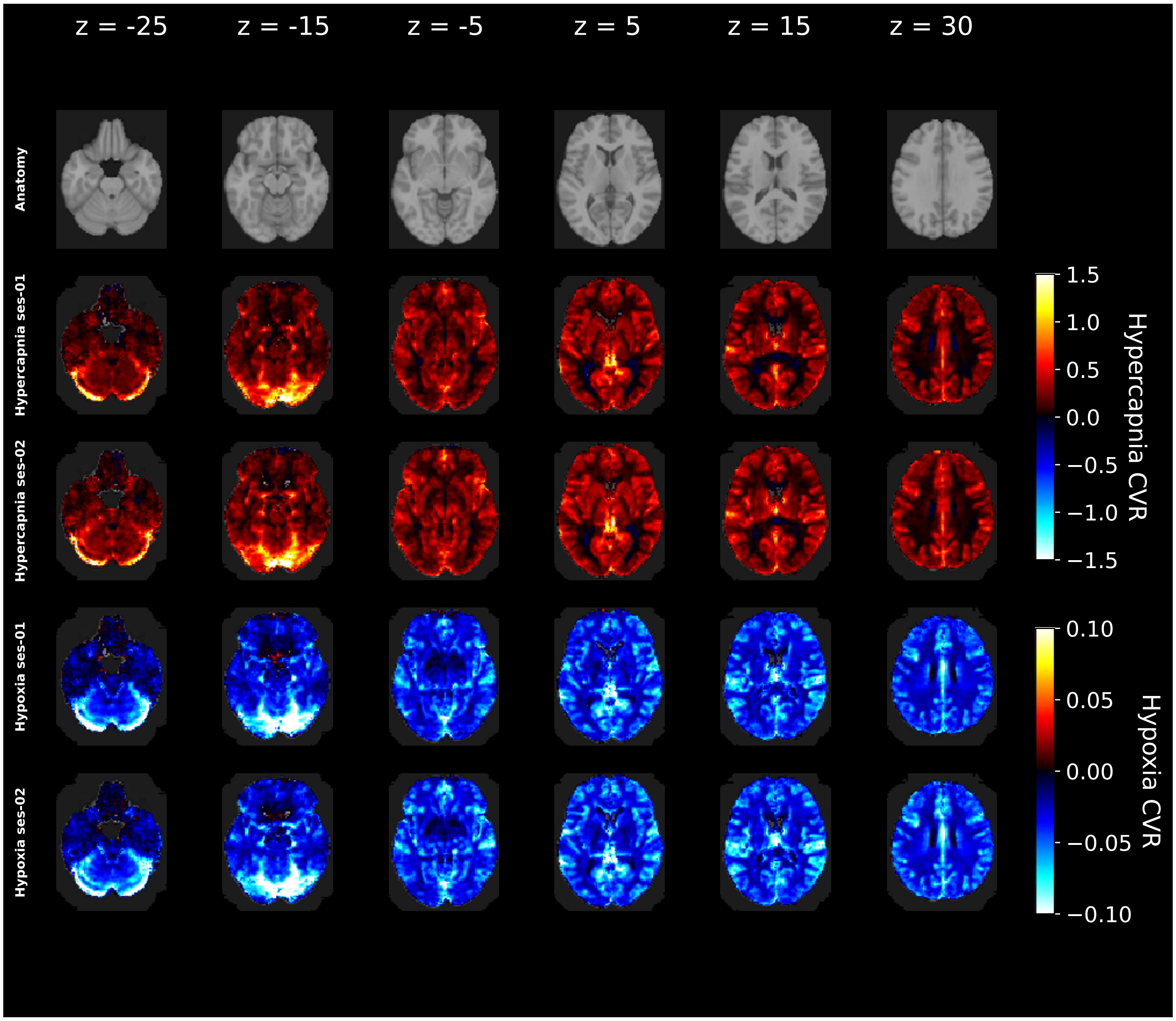


Figure S4: FIR GLM-derived group-mean voxelwise CVR maps for hypercapnia first session (top row), hypercapnia second session (second row), hypoxia first session (third row), and hypoxia second session (bottom row). $CVR_{HC}$ is expressed in % $\Delta$BOLD per mmHg $\Delta ETCO_2$; $CVR_{HO}$ is expressed in % $\Delta$BOLD per mmHg $\Delta ETO_2$.

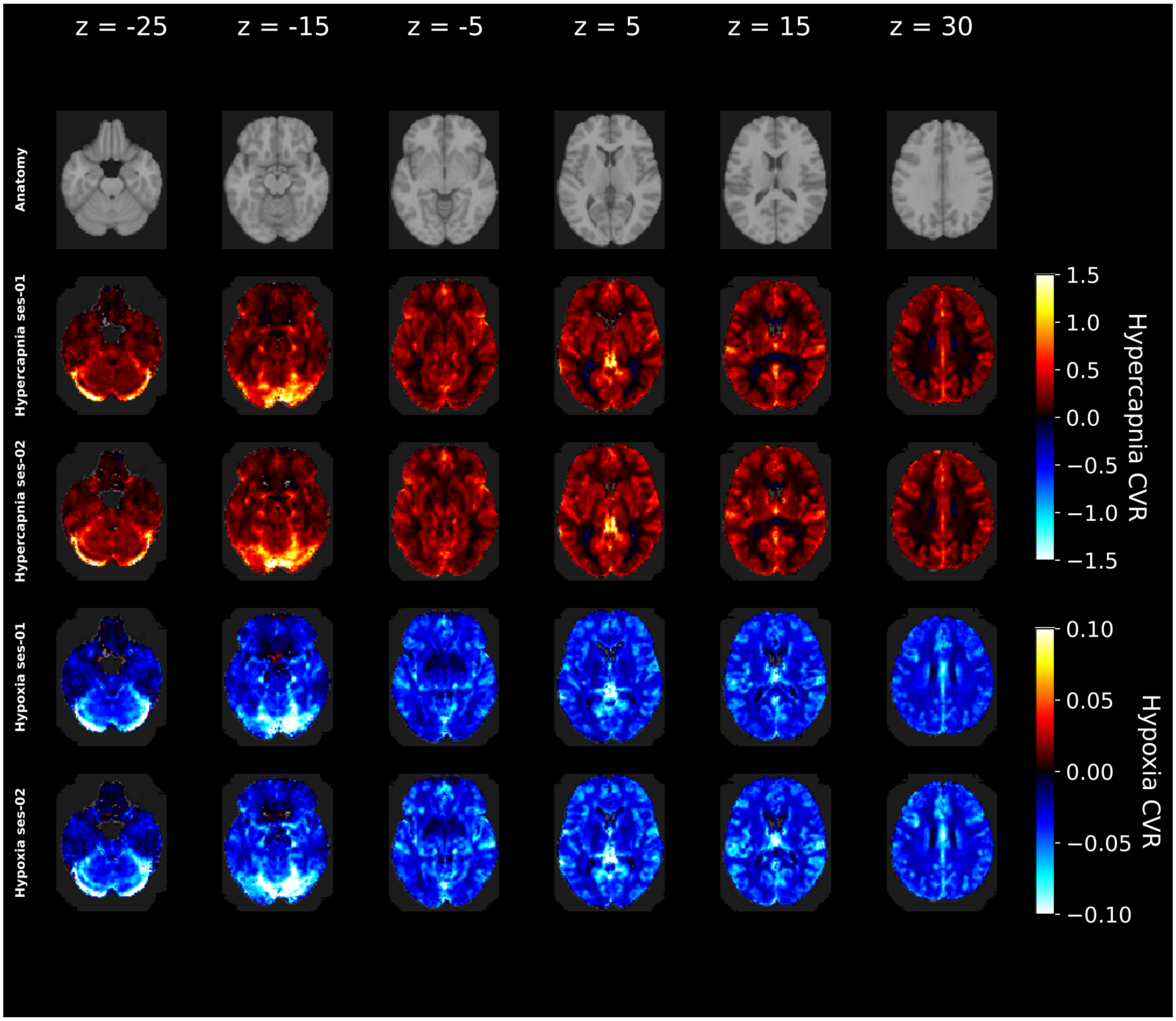


Figure S5: ET-trace GLM-derived group-mean voxelwise CVR maps for hypercapnia first session (top row), hypercapnia second session (second row), hypoxia first session (third row), and hypoxia second session (bottom row). $CVR_{HC}$ is expressed in % $\Delta$BOLD per mmHg $\Delta ETCO_2$; $CVR_{HO}$ is expressed in % $\Delta$BOLD per mmHg $\Delta ETO_2$.

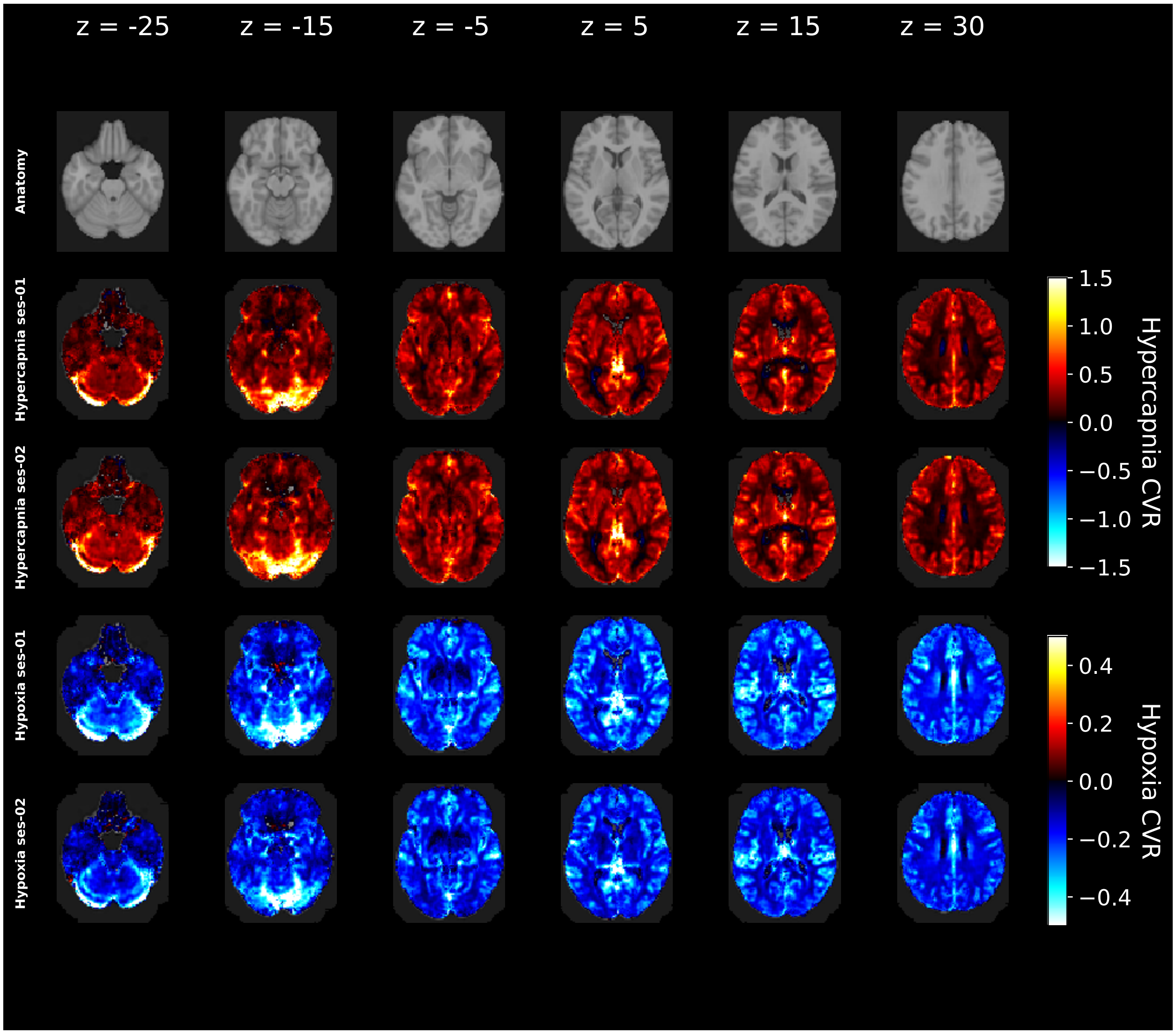


Figure S6: $SpO_2$-GLM-derived group-mean voxelwise CVR maps for hypercapnia first session (top row), hypercapnia second session (second row), hypoxia first session (third row), and hypoxia second session (bottom row). $CVR_{HC}$ is expressed in % ΔBOLD per mmHg $\Delta ETCO_2$; $CVR_{HO}$ is expressed in % ΔBOLD per Δ% $SpO_2$.

### Supplemental Figures: FIR-derived %ΔBOLD reproducibility (no gas-dose normalization)

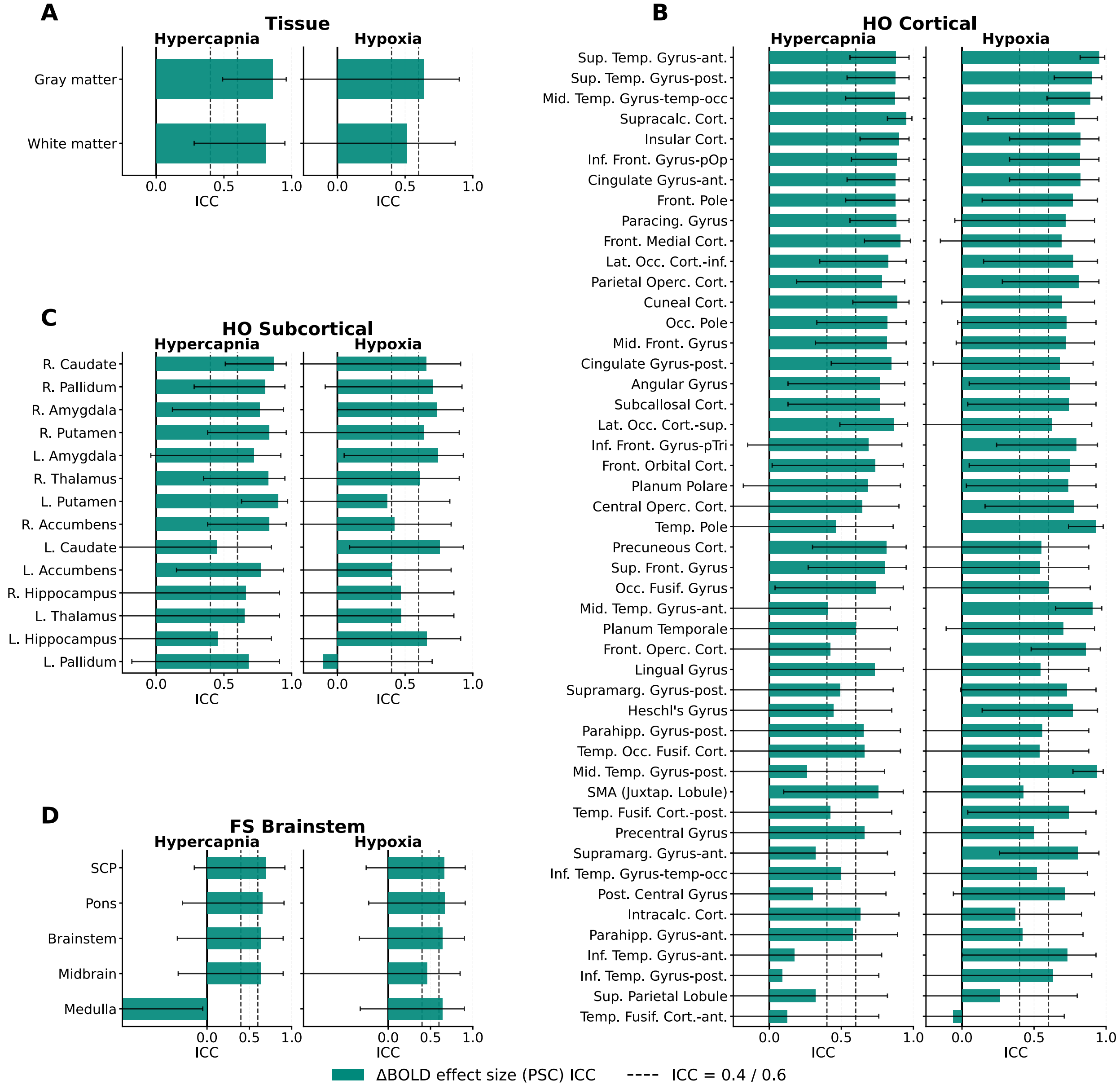


Figure S7: ROI-wise ICC of the FIR-derived %ΔBOLD (percent signal change per condition, not normalized to the delivered gas dose) across four atlases: (a) tissue compartments (gray matter, white matter), (b) Harvard–Oxford cortical ROIs, (c) Harvard–Oxford subcortical ROIs, and (d) FreeSurfer brainstem subdivisions. Within each panel, ROIs are ranked by mean ICC across hypercapnia and hypoxia; vertical reference lines mark ICC = 0.4 (fair) and 0.6 (good). Reproducibility of the unnormalized %ΔBOLD is generally acceptable across atlases, supporting the use of the FIR-derived %ΔBOLD as a normalization-free CVR-like measure for within-subject regional comparisons (main-text §4.3).

### Supplemental Figures: ROI-wise ICC for the FIR, ET-trace, and $SpO_2$ GLMs

The main text presents ROI-wise ICC results for the Severinghaus-derived $SaO_2$ GLM (main-text Figure 6), identified as the most cross-session-generalizable of the four models tested (main-text §3.3). The corresponding ROI-wise ICC panels for the FIR, ET-trace, and $SpO_2$ GLMs are shown below for direct comparison. All four models share the same 4-atlas layout — (a) tissue compartments (GM, WM), (b) Harvard–Oxford cortical ROIs, (c) Harvard–Oxford subcortical ROIs, and (d) FreeSurfer brainstem subdivisions — with vertical reference lines at ICC = 0.4 (fair) and 0.6 (good).

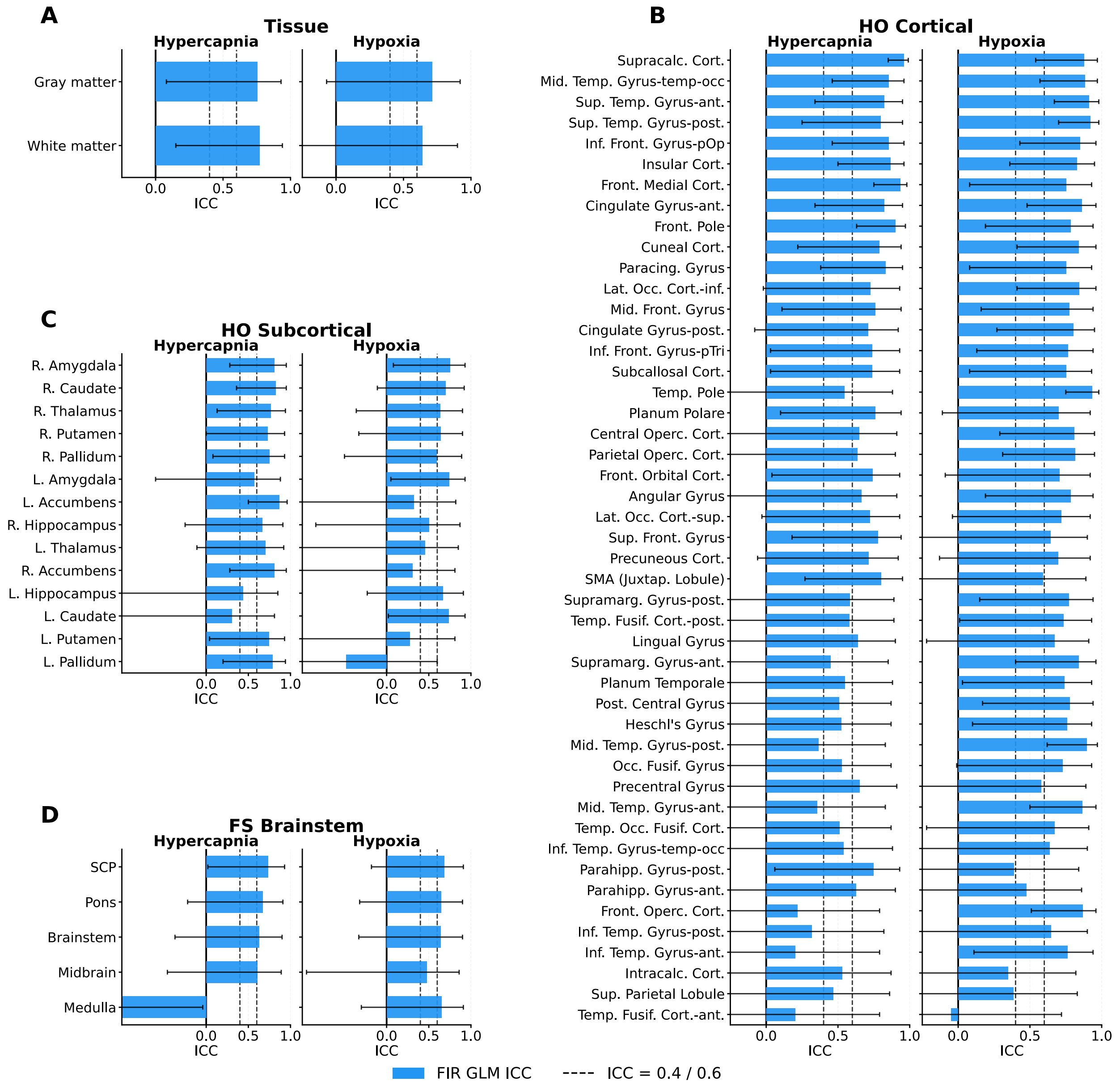


Figure S8: ROI-wise ICC for the FIR GLM (CVR normalized to $\Delta ETCO_2$ / $\Delta ETO_2$) across the same four atlases as main-text Figure 6.

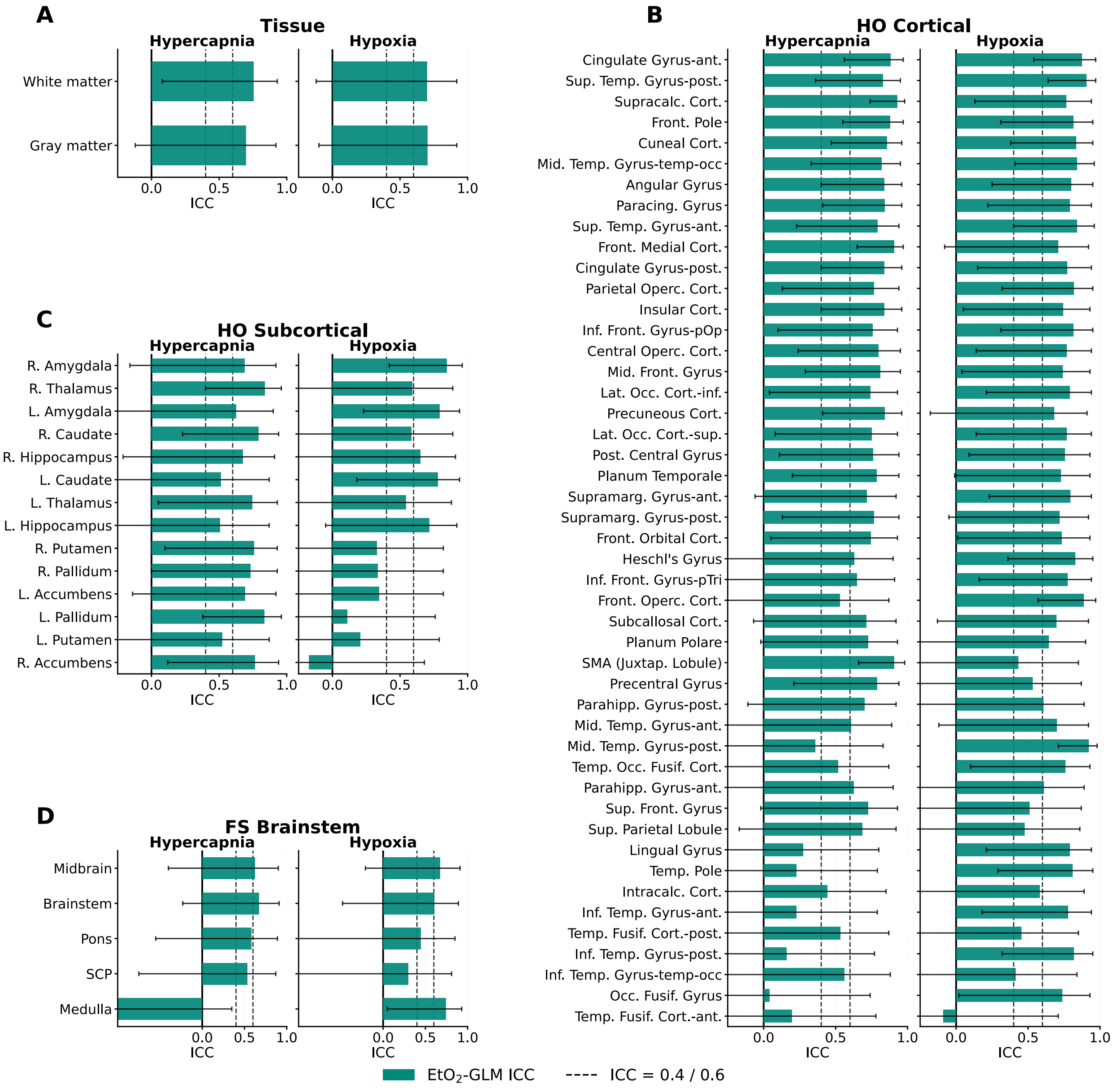


Figure S9: ROI-wise ICC for the canonical end-tidal-trace GLM ($ETCO_2$ / $ETO_2$ regressors) across the same four atlases as main-text Figure 6.

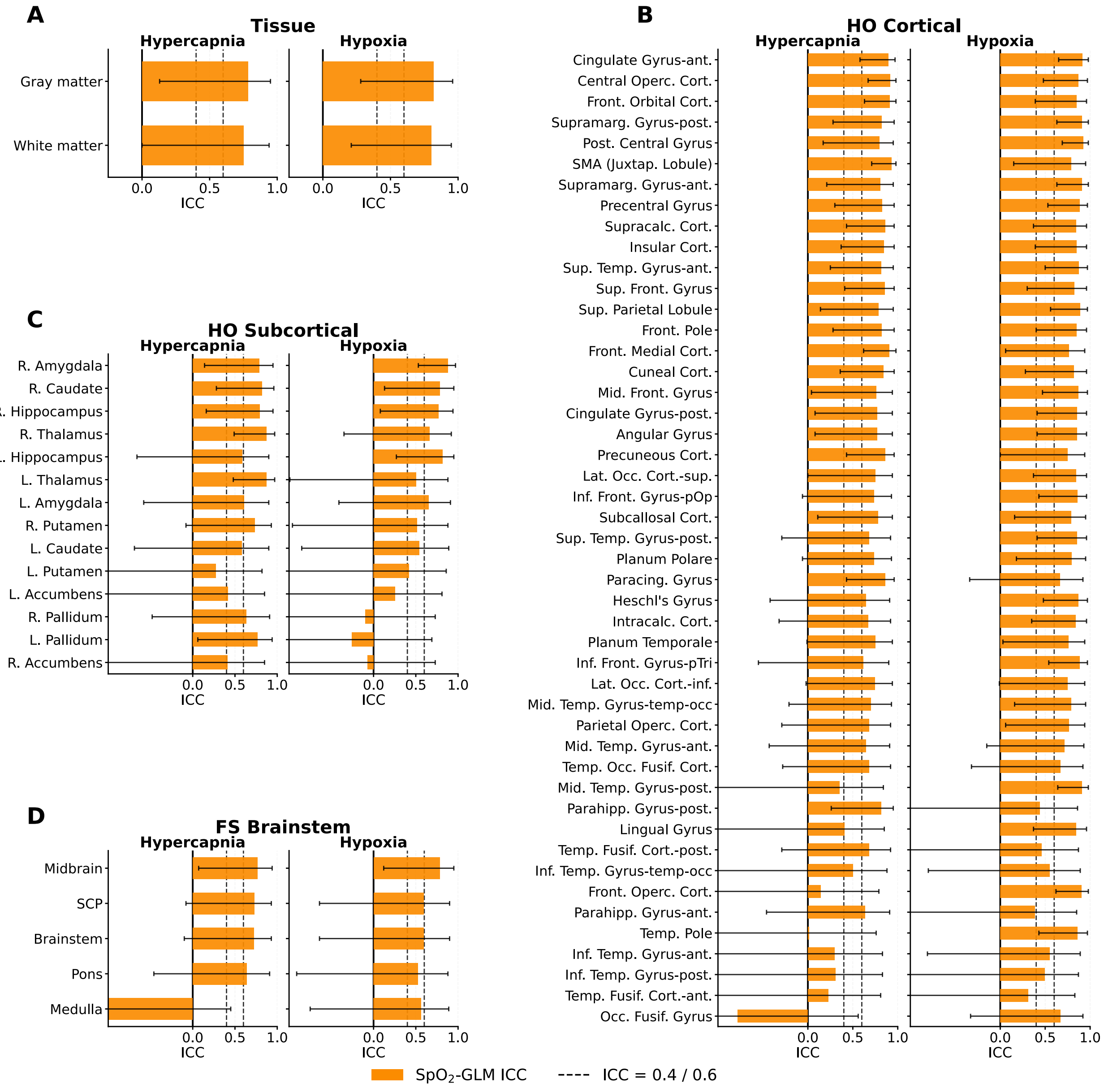


Figure S10: ROI-wise ICC for the pulse-oximetry $SpO_2$-based GLM across the same four atlases as main-text Figure 6.
